# Source-specific exposure and burden of disease attributable to volatile organic compounds (VOCs) in China’s residences

**DOI:** 10.1101/2025.08.25.25333590

**Authors:** Ningrui Liu, Chun-Sheng Huang, Yihui Yin, Xilei Dai, Jingjing Pei, Junjie Liu, Zhuohui Zhao, Yinping Zhang, Timothy Larson, Edmund Seto, Elena Austin

## Abstract

High-level exposure to indoor air pollutants (IAPs), including volatile organic compounds (VOCs), has substantially contributed to the burden of disease in China over the past two decades. However, the source contributions to the indoor VOC-related health burden remain unknown. This study utilized a novel approach based on positive matrix factorization (PMF) of indoor multipollutant data to estimate the source-specific residential VOC concentrations and associated burden of disease. Indoor concentrations of 39 VOCs were collected repeatedly in different seasons from 2016 to 2017 in 249 residences across nine cities in China. In 2017, the disability-adjusted life years (DALYs) attributable to residential VOC exposure across nine provinces in China reached 134.2 (95% UI: 65.7 – 225.0) per 100,000, resulting in financial costs of 28.1 (13.8 – 47.1) billion CNY. Contributions to indoor VOC concentrations from six indoor sources and three outdoor sources were derived by PMF. The top three sources, i.e., wood building materials and furniture, outdoor vehicle exhaust, and cooking and indoor combustion, accounted for 42.7%, 25.9%, and 11.0% of the VOC-attributable DALYs, which suggests prioritizing controlling these sources in China. This approach can be extended to other IAPs and provide fundamental data for future cost-benefit analysis of source control interventions.

**TOC Art:** 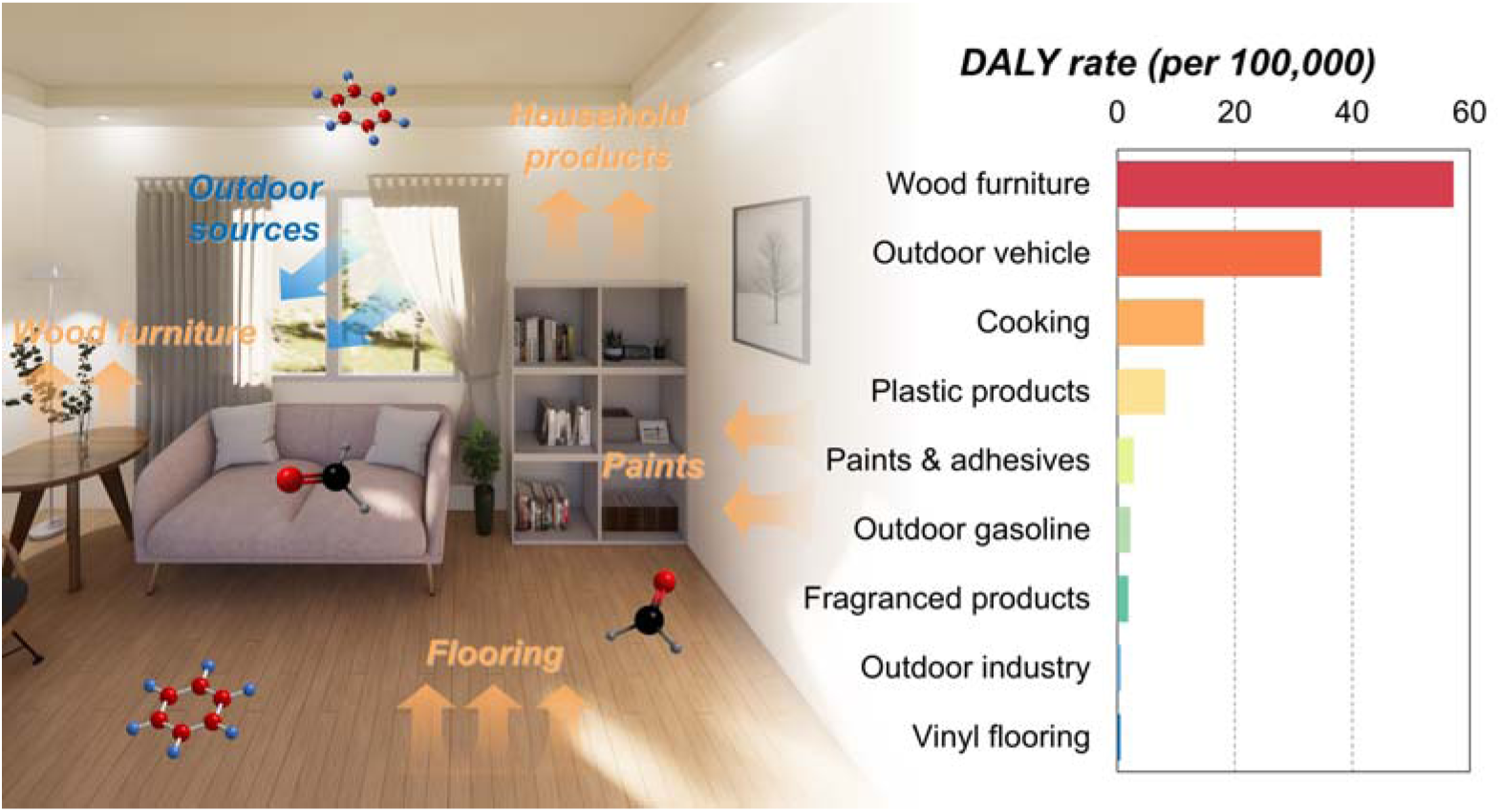

**Synopsis:** This novel method leverages multi-seasonal and multi-room residential VOC measurements to identify emission sources, quantify source-specific exposure concentrations, and estimate source-specific health burden, thus prioritizing the sources needing control.

## 1 Introduction

The Global Burden of Diseases Study (GBD) in 2021 found that air pollution is the third leading risk factor in China, accounting for 11.9% of total disability-adjusted life years (DALYs). ^1^ Since people spend more than 80% of their lifetime indoors, ^2, 3^ more attention should be paid to the health impacts of indoor air pollution. ^4, 5^ The attributable DALYs of ten indoor air pollutants (IAPs), such as fine particulate matter (PM_2.5_), nitrogen dioxide (NO_2_), and formaldehyde, in China were found to be 10-21% higher than those of outdoor air pollution during the past two decades. ^6^ Therefore, it is necessary to identify and control indoor air pollution sources to reduce their corresponding health burden.

The control strategies for indoor air pollution usually include source emission reduction, ventilation, and air purification. ^7^ Compared to source control, ventilation and air purification are usually easier to implement and are commonly used. Health burden-oriented optimized ventilation and air purification strategies for IAPs have been proposed in recent years, considering their health benefits or trade-offs with control costs. ^8–11^ However, long-term mechanical ventilation and air purification can incur financial costs and lead to additional energy consumption.

In contrast, source control is often more effective because it addresses pollution through eliminating the root source. ^12^ Unfortunately, it is still challenging to quantify the health benefits of source control interventions because of the complicated relationships between source emission and exposure levels. Based on survey data and the principle of mass conservation, Hu et al. conducted a simulation study to estimate the contribution of different sources (i.e., outdoor source, cooking, and second-hand smoking) to the health burden for residential PM_2.5_ and NO_2_. ^13, 14^ Then the health benefits of several source control interventions were obtained, including banning smoking, replacing gas stoves with electric stoves for cooking, and reducing outdoor concentrations. ^13, 14^ Besides PM_2.5_ and NO_2_, indoor volatile organic compounds (VOCs), such as formaldehyde and benzene, can also rank among the top 5 contributors to DALYs in many provinces in China. ^6^ However, residential VOC sources are more complex, including wood products, flooring, textiles, paint, furnishings, cleaning products, cooking, and outdoor infiltration, ^15, 16^ which presents challenges for clearly understanding the source contributions to VOC concentrations. Addressing these challenges would inform policy and decision-making in identifying and prioritizing source(s) for targeted interventions to improve health.

Previous studies have used two different approaches to understand the contributions of various indoor VOC sources. The first approach has been to develop source emission models (from source to exposure). A series of mass transfer models have been proposed to describe the VOC emissions from indoor materials, relying on mass-transfer mechanistic parameters of materials (e.g., initial emittable concentration, diffusion coefficient, and partitioning coefficient) and building characteristic parameters (e.g., air exchange rate). ^16–18^ These models have required a priori quantification of these parameters via laboratory experiments. ^18–20^ However, when applying these models in realistic indoor environments to estimate the source contributions to VOC levels, there has been several challenges. First, it is difficult to measure the mechanistic parameters for each existing indoor material separately, which is very time-consuming and expensive. Second, these mechanistic parameters are also dependent on temperature and humidity which is time-varying indoors, ^21–23^ making emission models more complicated. Therefore, the source emission models have been only feasible for assessing the impact of a few specified materials on indoor VOC concentrations, and not practical for the goal of screening large quantities of indoor materials to prioritize those for control.

The second approach has been to develop receptor models based on the measured multi-pollutant indoor concentrations in order to trace back from exposure to source. Receptor models are frequently used for source apportionment (SA) of ambient air pollution, especially ambient particulate matter based on particle size distribution and chemical composition. ^24, 25^ The commonly used receptor models include principal component analysis (PCA), positive matrix factorization (PMF), and other unsupervised statistical models. ^24, 25^ Recently, this approach has been applied to apportionment of IAPs. ^26^ Many studies have leveraged metals and polycyclic aromatic hydrocarbons (PAHs) to identify multiple sources of indoor PM_2.5_ (e.g., smoking, cooking, resuspension, and outdoor traffic emission) and then estimate their source contributions. ^27–37^ Nevertheless, such studies on indoor VOC SA are very limited. ^38–45^ Bari et al. conducted one such study, which measured over 100 VOCs for a continuous 24-hour period during the winter across 50 homes in Edmonton, Canada. Through PMF analysis, a total of 13 sources were derived, including nine indoor sources and four outdoor sources, and source contributions to total VOC (TVOC) concentrations were presented. ^38^ Limitations still exist among these VOC studies. First, most studies have obtained the short-term average VOC concentrations with limited repeated measurements (≤ 4) in one season, which may not reflect the long-term VOC concentrations related to population health. ^38–42, 44, 45^ Second, studies have largely focused on the source contributions to TVOC concentrations, but the health effects of different VOCs can differ considerably. ^38, 39, 44^ Source-specific exposure assessment for each VOC is required. Third, a few studies have estimated the health risks of VOC exposures, but none of them have provided the source-specific health burden, which is useful for source control intervention. ^39, 46^

Therefore, this study proposes an integrated approach to address the above research gaps on SA of indoor VOCs. Using residential VOC measurements across different seasons in nine cities in China, this study aims to (1) identify the sources of indoor VOCs, (2) quantify the source-specific long-term exposure concentrations of multiple VOCs, and (3) evaluate and rank the source-specific burden of disease attributable to indoor VOCs.

## 2 Methods

### 2.1 Data collection

The field measurement of indoor VOCs was conducted in 249 residences across nine cities in China from November 2016 to November 2017. ^47, 48^ We selected nine cities across diverse geographic and climatic regions of China, i.e., Tianjin, Shenyang, Shanghai, Changsha, Wuhan, Chongqing, Kunming, Xi’an, and Urumqi, to enhance spatial representativeness of residential indoor air conditions (shown in Supporting Information (SI) 1 **Figure S1**). A total of 1290 samples were collected, which covered different seasons in a year and different rooms (i.e., bedrooms, kitchens, and living rooms) for each residence. According to the national standard of indoor air quality in China (GB/T 18883), all windows and doors were closed for at least 12 hours prior to the indoor VOC measurements. Briefly, formaldehyde was collected at 0.5 L/min for 20 min and absorbed into the 3-methyl-2-benzothiazolinone hydrazine (MBTH) aqueous solution. Then the formaldehyde concentrations were measured by the spectrophotometer using visible adsorption at 630 nm. Other VOCs were collected at 0.5 L/min for 20 min into the Tenax-TA tubes. They were then desorbed by a thermal desorber (TD) and quantified by gas chromatography mass spectrometry (GCMS) method. Detailed sampling, chemical analysis, and quality assurance and control (QA/QC) information were described in previous papers. ^47, 48^ Since this study directly used the previously published dataset and did not collect new data, University of Washington Human Subjects Division (HSD) determined that this study does not involve human subjects (IRB ID: STUDY00023774), and review and approval by the University of Washington Institutional Review Board (IRB) is not required.

### 2.2 Source-specific exposure assessment

As a widely used receptor model, PMF was selected for source apportionment of indoor VOCs in this study. We applied PMF to resolve the observed concentration matrix (X) into two non-negative matrices: a factor profile matrix (F) and a factor contribution matrix (G), such that X ≈ G × F. ^49^ The model is

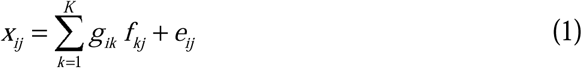

where *x_ij_* is the concentration of species *j* in sample *i*; *f_kj_* is the species profile of factor *k*, i.e., the concentration of species *j* in factor *k*; *g_ik_* is the contribution of factor *k* in sample *i*; *e_ij_* is the residual; and *K* is the number of factors. The factor profile and contribution matrices were derived by minimizing the squared uncertainty-scaled residuals under the constraints that factor profiles and contributions should be non-negative. ^49^

Before running the PMF model, the measured VOC concentrations need to be pre-processed. First, VOCs were included in the subsequent source apportionment if its detection ratio (proportion of measurements over detection limit) was higher than 25% because too many species concentrations under the detection limit cannot provide useful information on the variation of that species across different samples. A total of 39 VOCs were finally selected, which are listed in **Table S1**. Second, according to guidelines for PMF, the concentration *x_ij_* and the corresponding uncertainty σ*_ij_* were modified by Equation (2) and (3) below, respectively. ^49^

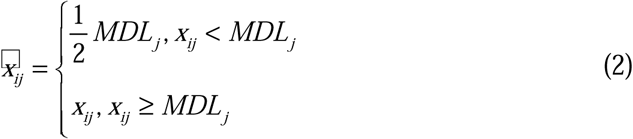

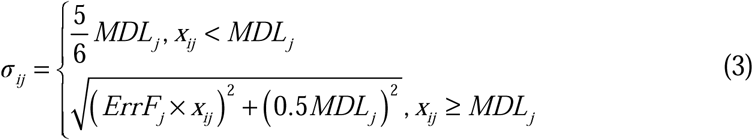

where *MDL_j_* is the method detection limit for species *j* (i.e., 5.6 μg/m^3^ for formaldehyde and 0.5 μg/m^3^ for other VOCs); and *ErrF_j_* is the error fraction of the measured concentrations of species *j* (i.e., 10%). ^47, 48^ Third, a few missing data were imputed to take full advantage of all collected samples, ^49^ which is described in **SI1 Section S1**.

We performed all PMF analysis using the EPA PMF 5.0 software in a spatiotemporal context. All 1290 samples from 249 residences in nine cities across four different seasons were included in one PMF model. In addition to 39 VOCs, TVOC concentrations were also included, which was the sum of formaldehyde concentrations and all detected VOC concentrations in the Tenax-TA tubes. In this study, based on signal-to-noise (S/N) ratio, all 39 included VOCs were “strong” species. ^49, 50^ TVOCs were categorized as “weak” as it was regarded as the total variable.

We tested models with 3–10 factors and selected the optimal solution based on the maximum individual column mean (IM) and standard deviation (IS) of the scaled residuals. ^51–54^ Factor interpretability was further improved by reclassifying poorly-fit ‘strong’ species to ‘weak’. ^55, 56^ We finally selected nine factors in this study. Detailed modeling methods are described in **SI1 Section S2.1**.

After the PMF modeling, we applied several diagnostic approaches to ensure the reliability of our PMF results. ^57, 58^ First, we used bootstrap methods to check if the spatiotemporal constant factor profile assumption was appropriate and the resulting solutions were robust or not. Robustness was evaluated using 100 block bootstraps stratified by city. Factor stability was confirmed when >80% of bootstrap runs mapped cleanly to base case profiles. Second, rotation ambiguity was assessed via G-space and displacement (DISP) analysis; the absence of oblique edges and factor swaps indicated stable solutions. If there were no obvious oblique edges in the G-space plot and no factor swaps in the DISP analysis, there were no significant rotational ambiguity. Detailed diagnosis methods are shown in **SI1 Section S2.2**.

Finally, we employed several external validation approaches to help interpret the possible VOC emission sources for each PMF factor. ^50^ First, VOC tracers in each factor profile played a critical role in source identification and were compared with previous emission characteristics or SA studies. Second, we calculated the difference in factor contributions between paired samples according to different seasons (summer vs winter), different room types (kitchens vs non-kitchen rooms (i.e., bedrooms and living rooms)), and different renovation times (within 1 year, 1-2 years, 2-5 years, and over 5 years). Then we obtained the associations between the contributions of each factor and these external variables to inform the factor interpretation. Third, ratios between different pollutants were applied, such as the ratio of toluene to benzene (T/B ratio).

Based on the above source apportionment, we estimated the average concentration of pollutant *j* attributable to source *k* in city *c* (denoted as *C_jk_*_,*c*_) as below. ^59, 60^

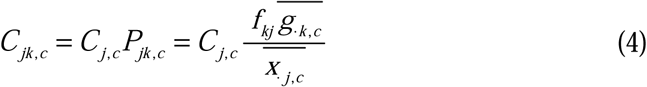

where *C_j_*_,*c*_ is the indoor concentration of pollutant *j* in city *c*, which used the previous modeling results in 2017 in Liu et al. for formaldehyde, benzene, and toluene, ^6^ or was equal to the measured concentrations in this study for other VOCs; *P_jk_*_,*c*_ is the percentage of contribution of source *k* in the total indoor concentrations of pollutant *j* in city *c*; *f_kj_* is the concentration of pollutant *j* in the profile of source *k*; 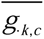 is the average contribution of source *k* in city *c*; and 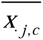 is the average concentration of pollutant *j* in city *c* measured in this study.

### 2.3 Source-specific attributable burden of disease

We estimated attributable health burden following the Global Burden of Disease (GBD) framework.^1^ Exposure-response (E-R) functions for formaldehyde, benzene, and toluene were drawn from recent meta-analyses; ^61, 62^ inhalation unit risks (URs) were sourced for VOCs with carcinogenicity evidence including formaldehyde, dichloromethane, 1,2-dichloroethane, acetaldehyde, ethylbenzene, styrene, and naphthalene. ^63–65^ Detailed E-R relationships are available in **SI1 Section S3.1**.

Population attributable fractions (PAFs) for pollutant *j* and outcome *d* in province *c* (denoted as *PAF_jd_*_,*c*_) were calculated using relative risks (Equation 5) or unit risks (Equation 6). ^1, 66–68^

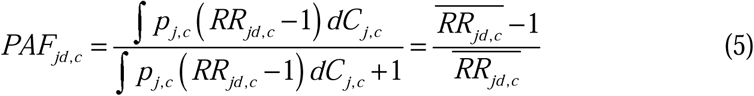

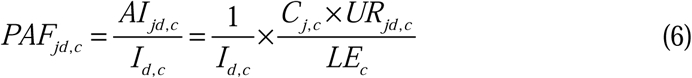

where *RR_jd_*_,*c*_ is the relative risk of outcome *d* for *C_j_*_,*c*_ exposure to pollutant *j* in province *c*; *p_j_*_,*c*_ is the population fraction for *C_j_*_,*c*_ exposure to pollutant *j* in province *c*; *AI_jd_*_,*c*_ is the attributable incidence, that is the annual cancer cases, of outcome *d* due to exposure to pollutant *j* in province *c*; *I_d_*_,*c*_ is the baseline incidence of outcome *d* in province *c* in 2017; *UR_jd_*_,*c*_ is the unit risk of outcome *d* for *C_j_*_,*c*_ exposure to pollutant *j* in province *c*; and *LE_c_*is the life expectancy in province *c* (details in **SI1 Section S3.2**). Then the source-specific PAF from source *k* for pollutant *j* and outcome *d* in province *c* (denoted as *PAF_kjd_*_,*c*_) was obtained using the proportional PAF approach as below. ^1^

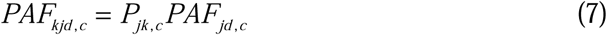

The burden of disease from source *k* for outcome *d* in province *c* (denoted as *DALY_kd_*_,*c*_) was first estimated, and then the total burden of disease attributable to source *k* in province *c* (denoted as *DALY_k_*_,*c*_) can be expressed by ^1^

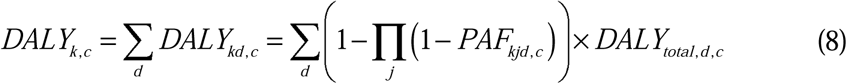

where *DALY_total_*_,*d*,*c*_ is the total burden of disease for outcome *d* in province *c* in 2017 (details in **SI1 Section S3.2**). ^69^

Based on the source-specific burden of disease, an adapted human capital method was finally applied to evaluate the source-specific financial costs according to gross domestic product (GDP) per capita in 2017. ^6, 70, 71^

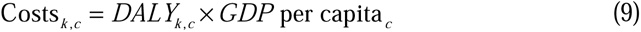

### 2.4 Uncertainty analysis

We estimated 95% uncertainty intervals (UIs) using a two-stage Monte Carlo simulation. ^6, 9, 14^ The first stage captured intra-population variability in VOC concentrations; the second stage incorporated uncertainties in exposure–response parameters and source contribution estimates (i.e., *P_jk_*_,*c*_ in Equation (4)) (details in **SI1 Section S3.3**). Except the source apportionment, all statistical analyses introduced in this section were performed with R V4.2.2.

## 3 Results and discussion

Summary statistics of residential concentrations of 39 included VOCs and TVOCs are shown in **Table S9**. The median (interquartile range, IQR) TVOC concentration was 371.3 (439.2) μg/m^3^, among which the three top VOCs were formaldehyde (64.6 μg/m^3^), toluene (11.9 μg/m^3^), and xylenes (9.3 μg/m^3^) according to indoor concentrations. Among four different seasons, summer had the highest TVOC level (599.8 μg/m^3^), while winter the lowest (301.4 μg/m^3^). For three room types, the highest and lowest TVOC median concentrations were found in bedrooms (399.1 μg/m^3^) and kitchens (288.9 μg/m^3^), respectively. The TVOC concentrations also differed between provinces, ranging from 183.0 μg/m^3^ in Hunan province to 662.1 μg/m^3^ in Liaoning province.

### 3.1 Source apportionment

A total of nine factors were identified from the PMF analysis. The factor profiles are shown in **Figure 1** and **Table S5**. The PMF solution was robust according to the block bootstrap method, and there was no significant rotational ambiguity based on the G-space plot and DISP analysis (details in **SI1 Section S2.2**). Among the nine factors, six factors were interpreted as indoor sources (Factors 1-6), while the other three factors were outdoor sources (Factors 7-9).

**Figure 1.**
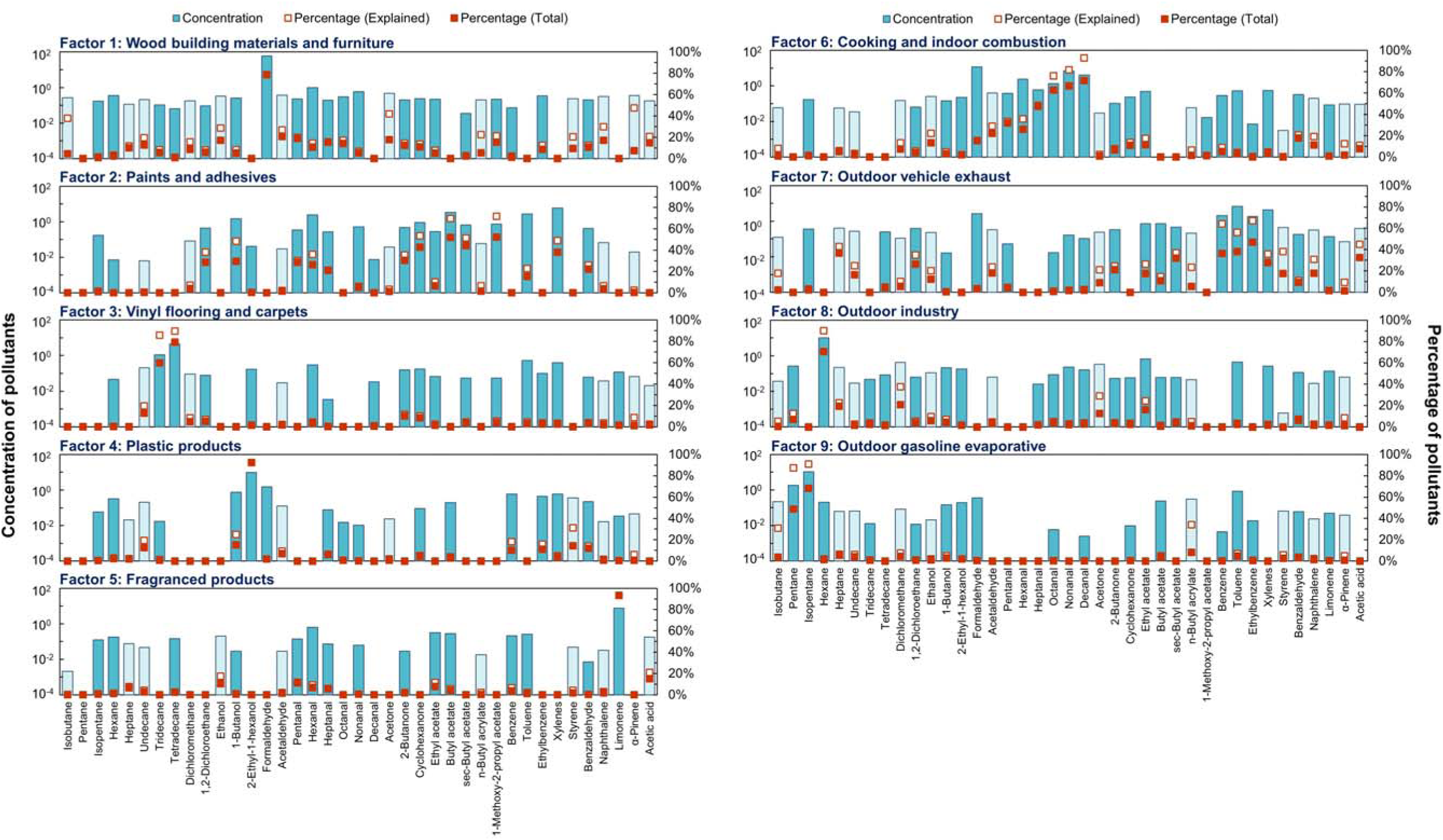
Profile of the nine PMF factors. Blue bars describe the pollutant concentrations in each factor, where dark blue and light blue refer to “strong” and “weak” species, respectively. Solid and hollow red points represent their percentage of the corresponding source contribution in the total pollutant concentrations and the sum of pollutant concentrations explained by the PMF model.

#### 3.1.1 Indoor sources

Factor 1 was considered as emissions from wood building materials and furniture. As can be seen from **Figure 1**, factor 1 was distinguished by formaldehyde, explaining over 70% of total formaldehyde concentrations. In addition, the percentage in explained concentrations was relatively high for acetone and α-pinene. Aldehydes were found in this factor, such as acetaldehyde and hexanal. Previous studies have found that emissions from wood building materials and furniture emission are enriched in formaldehyde, hexanal, acetaldehyde, and α-pinene, consistent with this factor profile. ^72–76^ Additionally, as shown in **Figure S5**, the contribution of factor 1 in summer was higher than that in winter. Higher temperature in summer usually increases the formaldehyde/VOC emissions from building materials, since higher temperature can increase the diffusion coefficient, help the residual formaldehyde trapped in the wood material overcome the bonding forces, as well as accelerate the hydrolysis of urea-formaldehyde (UF) adhesives to generate more formaldehyde. ^21, 22,76^ Higher humidity in summer can also promote these hydrolytic reactions. ^77^ **Figure S7** demonstrates that the factor contribution was higher in newly renovated residences than that in older residences. This agrees well with the reported association between renovation time and indoor formaldehyde concentrations in a systematic review. ^78^

Factor 2 was associated with paints and adhesives. This factor was characterized by butyl acetate, sec-butyl acetate, 1-methoxy-2-propyl acetate (PGMEA), toluene, xylenes, 2-butanone, cyclohexanone, 1,2-dichloroethane, 1-butanol, pentanal, hexanal, heptanal, and benzaldehyde. Toluene and xylenes were listed in the top 10 VOCs according to their contributions to emissions from both solvent-based and water-based architectural/furniture paints. ^79^ Butyl acetate and PGMEA were also found in solvent-based paints, while 1-butanol was abundant in water-based paints. ^80^ Additionally, several studies measured VOC concentrations emitted from solvents in furniture manufacturing factories, which identified ethyl acetate, butyl acetate, PGMEA, toluene, xylenes, ethylbenzene, cyclohexanone, butanone, dichloromethane, and 1,2-dichloroethane. ^81, 82^ The contribution of factor 2 was much higher in summer than that in winter (**Figure S5**), possibly due to higher emission rate from paints with higher temperature. The residences renovated within 1 year also had a higher factor contribution (**Figure S7**), supporting the idea that this factor is related to renovation activities.

Factor 3 may represent emissions from vinyl flooring and carpets. This factor was distinguished by tridecane and tetradecane. Previous studies suggest that vinyl flooring usually emits tridecane, tetradecane, pentadecane, and phenol. ^83–86^ Tridecane and tetradecane were also found in carpets. ^85–87^ **Figure S5** illustrates that the contribution of factor 3 was higher in summer than that in winter, which is consistent with higher VOC emission rate under higher temperatures, similar to factors 1 and 2. In addition, the factor contribution was much higher in residences renovated within 1 year than those renovated beyond 1 year (**Figure S7**), suggesting that factor 3 should be one of flooring materials used during the renovation.

Factor 4 was interpreted as emissions from plastic products. The major species in factor 4 were 2-ethyl-1-hexanol (accounting for 92.4%), 1-butanol, and formaldehyde. A comprehensive review summarized that the primary source of indoor 2-ethyl-1-hexanol was the hydrolysis of plasticizers and flooring adhesives, especially di(2=ethylhexyl) phthalate (DEHP) in polyvinyl chloride (PVC) products. ^88^ Similar emissions from hydrolysis reaction happen for 1-butanol, which is produced from di=n=butyl phthalate (DnBP). ^88^ A higher factor contribution was also observed in summer than that in winter (**Figure S5**), which agrees well with higher emission rate under higher temperatures.

Factor 5 was likely emissions from fragranced products. This factor was mainly comprised of limonene. Limonene is widely used in various household products with added fragrance, such as cleaning products, deodorizers, air fresheners, essential oils, and personal care products. ^89–91^ **Figure S6** shows that the factor contribution was higher in kitchens than that in non-kitchen rooms. That’s possibly because there is more household cleaning product usage in kitchens to wash dishes and remove the grease and grime on ovens and stoves. There is no significant difference in factor contributions between different seasons or different renovation times in **Figures S5** and **S7**, respectively.

Factor 6 was identified as cooking and indoor combustion. The factor profile was dominated by various aldehydes, including formaldehyde, acetaldehyde, pentanal, hexanal, heptanal, octanal, nonanal, and decanal. The cooking process, especially Chinese cooking, can generate a large amount of aldehydes, which can sometimes account for over 60% of TVOC emissions from cooking activities. ^48, 92–96^ The underlying mechanism is that, when the cooking oil and food are heated, the fatty acids (especially unsaturated ones) experience degradation and oxidization processes and are broken into small molecular compounds, like aldehydes. ^95, 97^ Coggon et al. pointed out that long-chain aldehydes (C > 6), such as octanal and nonanal, are more likely related to cooking oil fumes compared with short-chain aldehydes. ^92^ The T/B ratio of this factor was 1.9, which was close to 1.9-2.3 measured by Li et al. ^94^ **Figure S5** tells that the contribution of factor 6 was much higher in summer than in winter. Higher temperatures in summer may accelerate the degradation of fatty acids. More importantly, several studies revealed that ozone can react with the fatty acid constituents of cooking oil on various surfaces of kitchens and lead to secondary emissions of aldehydes. ^98–100^ Higher ozone concentrations in summer likely help produce more cooking-related secondary aldehyde emissions. Moreover, **Figure S6** shows that a higher contribution was observed in kitchens than that in non-kitchen rooms, further supporting that this factor is associated with cooking. **Figure S7** demonstrates that the newly renovated residences had a lower factor contribution than those old residences, ruling out the possibility of emissions from other building materials which may also emit aldehydes. However, we should note that other indoor combustion processes, such as smoking and incense/candle burning, can also emit abundant aldehydes and have similar profiles. ^101–103^ Therefore, we considered factor 6 as a combination of cooking and indoor combustion.

#### 3.1.2 Outdoor sources

We regarded factor 7 as outdoor vehicle exhaust. Key species identified in this factor included benzene, toluene, ethylbenzene, and xylenes (BTEX). We also observed ethyl acetate, butyl acetate, and 1,2-dichloroethane in this factor profile. Since both outdoor sources such as vehicle exhaust and indoor sources such as paints and adhesives generate BTEX, ^79, 104–107^ we interpreted factor 7 from several perspectives. First, **Figure S7** shows that the factor contribution was a bit lower in newly renovated residences than that in old residences, suggesting that factor 7 is probably not related to renovation materials such as paints and adhesives. Second, **Figure S5** suggests that the factor’s contribution was much higher in winter than in summer. If this factor was emission from paints and adhesives, it should have a higher contribution in summer, similar to factor 2. For outdoor vehicle exhaust, previous studies reveal that, in winter, reduced photochemical reactions due to less solar radiation, lower boundary layer height with more stagnant air, and higher BTEX emission rate due to incomplete combustion during the cold start of vehicles can contribute to higher ambient BTEX concentrations, ^108–111^ which infiltrate into indoors and lead to a higher contribution to indoor BTEX concentrations. Third, the T/B ratio of factor 7 was 3.3. Previous studies suggest that T/B ratio ranging from 1.5 to 4.3 reflects vehicle emission, while a higher T/B ratio refer to industrial emission or solvent evaporation. ^104^ Therefore, considering that all residences were located in urban areas, factor 7 may be outdoor vehicle exhaust.

Factor 8 was interpreted as outdoor industrial emissions. The profile was marked by the high levels of n-hexane, followed by considerable contributions from dichloromethane, acetone, heptane, and ethyl acetate. **Figure S7** demonstrates that there was no significant difference in factor contributions between newly renovated and old residences, suggesting its weak association with indoor renovation materials. Several studies measured the VOC profiles of various outdoor emission sources, and found that n-hexane was the major contributor to emissions from oil refinery and electronic manufacturing. ^112, 113^ Dichloromethane was also related to printing, auto manufacturing, plastic production, and the pharmaceutical industry. ^113–115^

Factor 9 was related to outdoor gasoline evaporation. The most abundant species in factor 9 were pentane and isopentane. As volatile short-chain alkanes, pentane and isopentane can be found in outdoor vehicle exhaust and gasoline evaporation. ^112^ The isopentane/n-pentane ratio of factor 9 was 6.0. Previous research shows that this ratio was 2.2-3.8 for vehicle emission and 1.8-4.6 for fuel evaporation. ^116^ Mo et al. also provided a ratio of 5.3 for gasoline vapor (China V). ^117^ The isopentane/toluene ratio of factor 9 was 12.8, which is consistent with 8.8-29.9 reported for headspace gasoline vapor emissions and higher than 0.3-1.9 reported for vehicle tailpipe emissions. ^118^

### 3.2 Source-specific exposure concentrations of residential VOCs

Based on source apportionment results, the source contributions to TVOC concentrations in China’s residences in 2017 were obtained, shown in **Figure 2** and **Table S10**. The top contributor to TVOC concentrations in 2017 was wood building materials and furniture, accounting for 16.4%. It was then followed by paints and adhesives (13.3%) and outdoor vehicle exhaust (12.5%). However, source contributions to TVOC concentrations had a heterogeneous spatial distribution (**Table S11**). While wood materials and paints ranked as the top two contributors in many provinces, outdoor vehicle exhaust ranked first in Hunan and Shaanxi provinces, and outdoor gasoline evaporative ranked first in Liaoning province. Cooking and indoor combustion also ranked as one of the top three contributors in five out of nine provinces.

**Figure 2.**
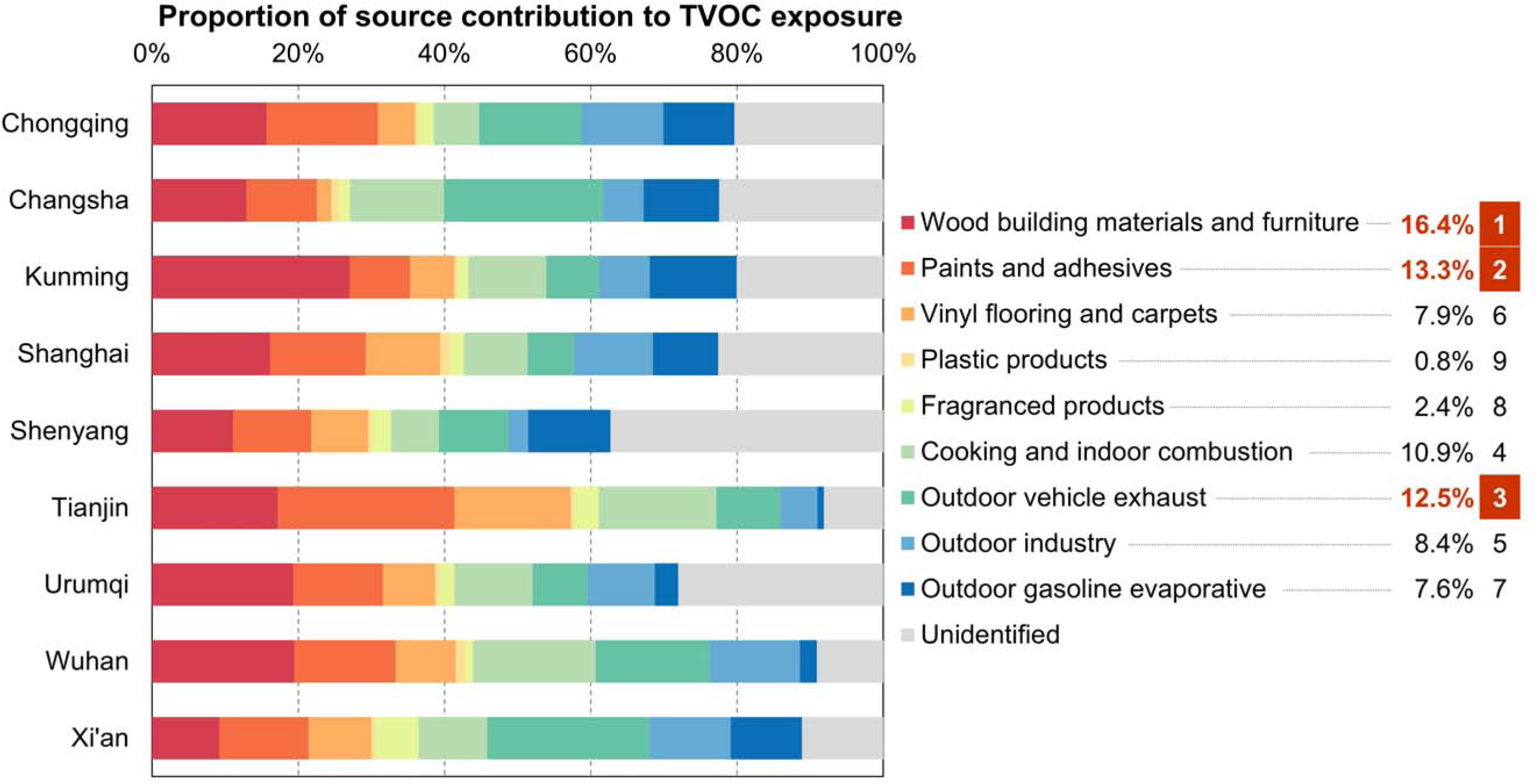
Proportion of source contributions to TVOC concentrations in residences of 9 cities in China in 2017. The figures on the right represent the average proportion and ranking of sources across the 9 cities.

In addition to TVOC concentrations, the source contributions to specific VOCs can differ. For instance, wood building materials and furniture contributed to 75.0% of formaldehyde concentration, followed by cooking and indoor combustion (15.9%) (**Figure S8**). By contrast, outdoor vehicle exhaust was the largest contributor to benzene concentration (43.1%) (**Figure S9**). Hexanal, detected in 88% of samples, was dominantly contributed by cooking and indoor combustion (31.7%), followed by paints (28.9%) and wood materials (11.4%) (**Figure S10**).

### 3.3 Source-specific burden of disease attributable to residential VOCs

In 2017, the burden of disease attributable to residential VOC exposure across nine provinces in China reached 134.2 (95% UI: 65.7 – 225.0) DALYs per 100,000, accounting for 0.47% (0.23 – 0.78) of total DALYs in 2017. The corresponding financial cost was 28.1 (13.8 – 47.1) billion CNY, which accounted for 0.13% (0.07 – 0.23) of total GDP in these 9 provinces. Detailed results are available in **Tables S12**-**S19**. **Figure 3** shows the source-specific DALY rates attributable to residential VOCs across the nine provinces in China in 2017. Wood building materials and furniture accounted for the largest share of the total VOC-related disease burden in 2017. Its attributable burden was 57.3 (39.2 – 79.6) DALYs per 100,000 in 2017, contributing 42.7% of total VOC health burden. The second and third source were outdoor vehicle exhaust and cooking and indoor combustion, which contributed 34.7 (8.8 – 81.9) and 14.7 (9.4 – 27.7) DALYs per 100,000 and accounted for 25.9% and 11.0% of VOC burden, respectively. While indoor sources contributed much of the VOC-related health burden, outdoor sources particularly vehicle exhaust, also had the substantial impact and warrant attention in research and mitigation efforts. Asthma (48.6%) and leukemia (41.9%) were the primary health outcomes associated with indoor VOC exposure, together accounting for over 90% of attributable DALYs. This reflects both the strength of the exposure-response evidence and the relatively high baseline incidence of these conditions. Other cancers based on URs, such as nasopharynx cancer and soft tissue sarcomas, resulted in very limited burden of disease due to residential VOC exposure.

**Figure 3.**
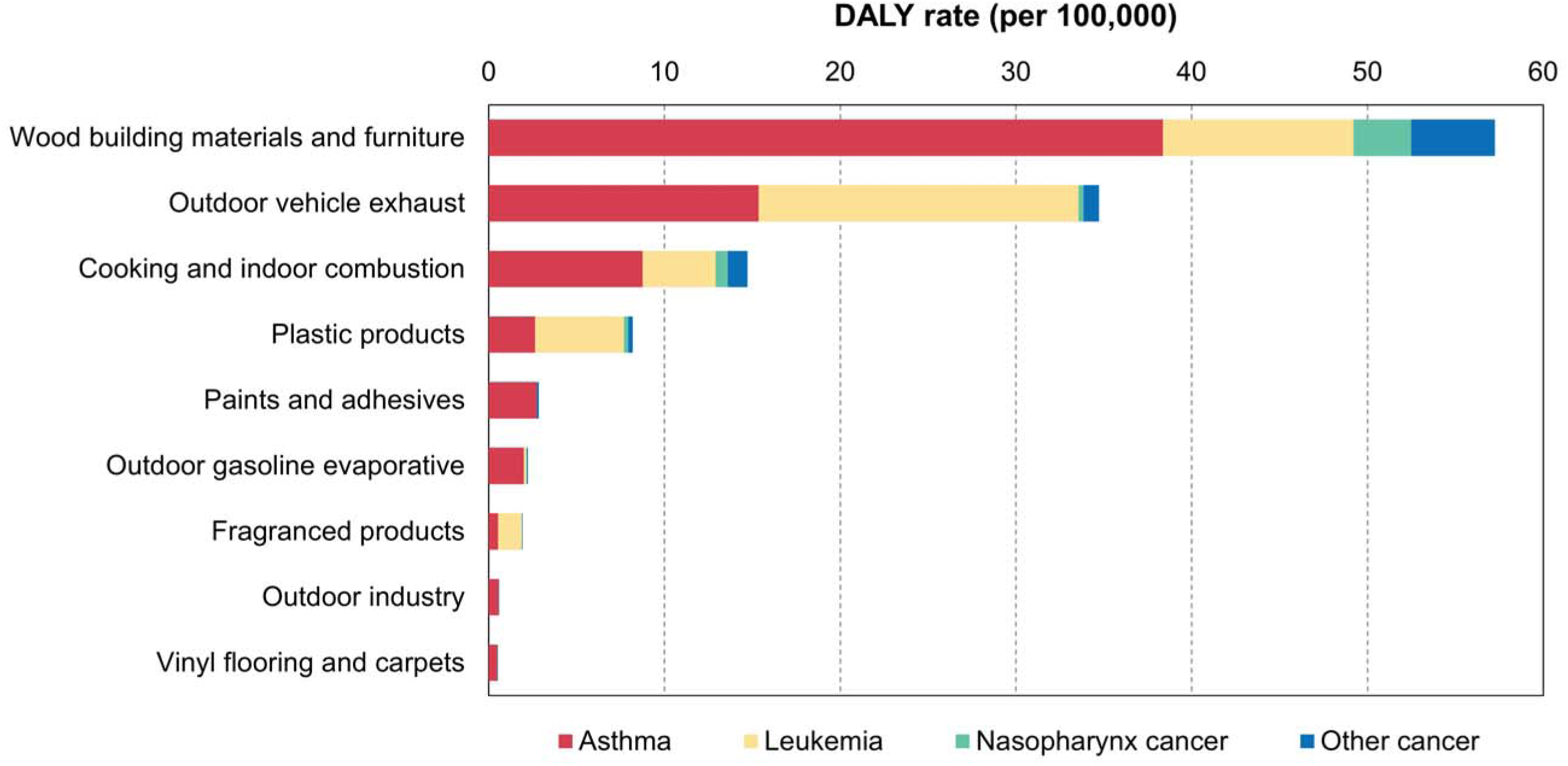
Source-specific DALY rates attributable to VOCs in China’s residences, by health outcome, 2017.

The national-level estimates indicate that wood building materials, infiltration of outdoor vehicle exhaust, and cooking and indoor combustion should be controlled in priority to reduce the VOC-attributable burden. However, the target sources may vary across different provinces. The provincial-level DALYs due to these VOC sources are provided in **Figure S11** and **Tables S12-S14**. As shown in **Table S20**, the top 3 sources remained the same as the national-level results in six provinces. In Chongqing, the burden from outdoor vehicle exhaust exceeded the wood building materials. In Hunan and Shanghai, plastic products were listed in the top 3 sources. This suggests that each province should have its own source control priorities.

We also focused on the spatial distribution of DALYs from the top sources at the national level across nine provinces (**Figure S12**). Yunnan and Tianjin province had the highest DALY proportion due to wood building materials, while outdoor vehicle exhaust contributed the largest DALY proportion in Yunnan and Chongqing provinces. It indicates that more efforts are required to control the corresponding sources in a few provinces.

We notice that the rankings were different between the source contribution to TVOC concentrations and that to burden of disease. For example, paints and adhesives ranked second based on contribution to TVOC concentrations but ranked only fifth according to burden of disease. This is because the strength and availability of E-R relationships differed considerably among various VOCs. Formaldehyde, benzene, and toluene contributed most of the attributable burden, resulting in relatively high health burden for sources abundant in these species, such as wood building materials and furniture. By contrast, many dominant species in the profile of paints and adhesives, such as butyl acetate, PGMEA, xylenes, and cyclohexanone, did not have available E-R relationships currently, thus providing a conservative estimate of this source’s health burden. Future epidemiological evidence may improve the estimation of residential VOC-attributable burdens and their rankings.

### 3.4 Comparison with previous studies

We found four studies which also used PMF models to apportion the sources of indoor VOCs. Three were in China and one was in Canada. ^38–41^ The study by Liu et al., which identified five factors from only seven VOC species, was excluded from detailed comparison. ^41^ The rational for this decision was that four out of five factors were dominated by only one species. The comparison of the source contributions to TVOC concentrations between this study and the other three studies is shown in **Table S7**. Building materials and furniture in this study (30.3% after scaling) had a similar contribution to the two Chinese studies (44.5% and 31.8%), but showed a higher contribution than Canada (8.0%). A similar pattern was observed for paints and adhesives. These findings suggest that the indoor pollution from building materials and paints in China was more severe than Canada. For cooking and indoor combustion as well as outdoor industry, the source contributions were similar between this study and previous studies. Compared to prior work, household consumer products contributed a small proportion of TVOC concentrations (4.0% after scaling) and was lower than other studies, especially the Canadian study (58.3%). Bari et al. found that one factor abundant in ethanol (100% detected) explained 44% of TVOC concentrations, ^38^ while the detection ratio of ethanol was only 46.6% in this study. The complexity of numerous household products may also lead to this difference. The contribution of outdoor vehicles was higher than previous studies. That’s possibly because the selected residences in this study were in and around the downtown area with high traffic volumes. The review of IAP SA studies also pointed out that the outdoor source contribution varied a lot among different studies due to regional conditions, building characteristics, and possible mixture of indoor and outdoor sources with similar profiles. ^26^

Besides these studies, other related studies applied PCA, factor analysis, or non-negative matrix factorization for indoor VOC SA. ^42–45, 119^ Different SA techniques may result in different source contributions, so we only focused on the identified sources. Norris et al. identified six sources in 20 residences in Shanghai, China, including vehicle emission, cooking, consumer products, solvents, wooden materials, and hexane sources, which was very similar to the sources identified in this study. ^42^ Guo et al. also found factors related to building materials, room freshener, wood products, household products, mothballs, and consumer products in 100 homes in Hong Kong, China. ^44^

We also compared the study design of these studies. Most of the above studies sampled the indoor VOCs for one to four continuous days. ^38–42, 44^ Those one-day measurements last for 24 hours, while other repeated measures only last for 1-2 hours. The exceptions were Rösch et al., sampling for 4 weeks, and Jia et al., measuring VOCs in two seasons. ^43, 45^ By contrast, this study collected the indoor VOC samples in multiple seasons across one year. In addition, most prior studies measured VOC concentrations in a single room like the living room, whereas this study collected data in living rooms, bedrooms, and kitchens, which can cover different indoor exposure scenarios of residents (discussed in **SI1 Section S4.3**).

A few studies measured both indoor and outdoor VOC concentrations simultaneously. Bari et al. performed SA for indoor and outdoor VOCs separately. ^38^ Huang et al. calculated the indoor/outdoor (I/O) ratios for all VOCs, helping determine if the VOC was dominantly from indoors or outdoors. ^39^ Norris et al. combined both indoor and outdoor data into one PMF model, and then compared the factor contribution in indoor and outdoor samples to identify whether each factor represented an indoor or outdoor source. ^42^ Most studies additionally collected questionnaires to provide source-related information and help interpret the factors. Future research can integrate the strengths of above study designs to improve the SA model performance and interpretability.

Finally, we compared the VOC-attributable burden in this study with other household risk factors in GBD. In 2017, the DALY rate of residential VOCs (134.2 per 100,000) was lower than household PM_2.5_ from solid fuels (750.1), secondhand smoke (723.6, both outdoor and indoor), and lead exposure (615.5, both outdoor and indoor), but higher than residential radon (44.3) and unsafe water, sanitation, and handwashing (22.9) in China. ^1^ Additionally, the DALYs of residential VOCs were much higher than those of occupational VOC exposure in China, which were only 9.8 per 100,000 for occupational formaldehyde, benzene, trichloroethylene, and PAHs. This is because, although industrial workers may experience much higher peak VOC exposures than residents, a much larger population is exposed to residential VOCs over significantly longer durations. Therefore, residential VOC control should continue to be emphasized by policy makers to improve population health in the future.

### 3.5 Strengths and limitations of this study

To our best knowledge, this is the first study to assess the source-specific burden of disease attributable to indoor VOCs in residences based on measured multi-pollutant concentrations. Instead of only ranking the pollutants, this study ranked the residential VOC sources based on health burden, which is helpful to prioritize the sources for control. Additionally, this study applied the PMF model to multi-seasonal and multi-room measurements. Besides, we also used a series of external validation approaches to interpret and ensure the reliability of the PMF results, including comparing the VOC markers with literature, comparing the difference of factor contributions in various subgroups such as seasons, and calculating ratios of different VOCs in the factor profile.

However, there are still limitations in this study. First, there is potential bias in spatial coverage of field measurements. Because measurements were restricted to urban residences, the results may not fully represent exposure conditions in rural areas, where building materials and ventilation practices may differ substantially. For each seasonal measurement, the 20-min sampling period was not so representative. Continuous sampling (e.g., 24 hours) with sufficient temporal resolution helps obtain more accurate source information. Second, considering the complexity of indoor VOC sources, incorporating additional external information can be helpful. More typical VOC/SVOC tracers for common sources should be added, such as nicotine for smoking, ^120^ and palmitic acid and cholesterol for cooking. ^121–123^ This study also lacked the questionnaire data except the building renovation time to help the factor interpretation, such as the materials/furniture used in residences, the cooking habit, and the presence of smokers. Outdoor samples can simultaneously be collected in future studies, which can be combined with indoor samples into the PMF model to tackle the challenge of identifying outdoor VOC sources. Third, pooling data across cities and seasons into a single PMF model allowed for generalizable source identification but may have masked city-specific profiles. ^50^ In this study, this concern should be mild as evidenced by the robust block bootstrap and DISP analysis. However, a few VOCs in some cities were not well explained by the current nine factors (details in **SI1 Section S4.4**), suggesting that there are potential city-specific factor profile variation or unidentified special sources. Fourth, the exposure-response relationships for some VOCs were still lacking, thus underestimating the source-specific VOC-related health burden. ^6, 61^ Most of the VOC-related epidemiological evidence was based on case-control and cross-sectional studies, ^6, 61^ and some cancer unit risk were obtained from animal toxicological evidence, which may bring potential bias in risk assessment. Some indoor sources, such as cooking and indoor combustion, can emit VOCs and other pollutants like PM_2.5_ at the same time, so estimating source-specific total health impacts is required in future studies.

In summary, the novel approach in this study can offer valuable insights on the source contributions to residential VOC concentrations and corresponding health burden. Wood building materials and furniture, outdoor vehicle exhaust, and cooking and indoor combustion were estimated to be the top three contributors to VOC-related health burden in China in 2017, suggesting their source control in priority. Our approach can be extended to other IAPs and other areas. Future studies can further perform the benefit-cost analysis of related source control interventions.

## Supporting information

Determination letter of UW HSD

## Data Availability

All data produced in the present study are available upon reasonable request to the authors

## Acknowledgements

This research was supported by The National Key Project of the Ministry of Science and Technology, China (2023YFC3708403), The New Chongqing YC Project (CSTB2024YCJH-KYXM0088), and The National Key Project of the Ministry of Science and Technology, China (Grant No. 2016YFC0700500).

## Notes

### Competing Interest Statement

The authors have declared no competing interest.

### Funding Statement

This study was funded by The National Key Project of the Ministry of Science and Technology, China (2023YFC3708403), The New Chongqing YC Project (CSTB2024YCJH-KYXM0088), and The National Key Project of the Ministry of Science and Technology, China (Grant No. 2016YFC0700500).

### Author Declarations

Human Subjects Division (HSD) of University of Washington (UW) waived ethical approval for this work. The UW HSD determined that this study does not involve human subjects (IRB ID: STUDY00023774), and review and approval by the University of Washington Institutional Review Board (IRB) is not required.

## References

1. GBD Risk Factors Collaborators, Global burden and strength of evidence for 88 risk factors in 204 countries and 811 subnational locations, 1990-2021: a systematic analysis for the Global Burden of Disease Study 2021. Lancet 2024, *403*, (10440), 2162–2203.

2. Duan, X., Exposure factors handbook of Chinese population. China Environment Publishing Group: Beijing, China, 2013.

3. Klepeis, N. E.; Nelson, W. C.; Ott, W. R.; Robinson, J. P.; Tsang, A. M.; Switzer, P.; Behar, J. V.; Hern, S. C.; Engelmann, W. H., The National Human Activity Pattern Survey (NHAPS): a resource for assessing exposure to environmental pollutants. Journal of Exposure Analysis and Environmental Epidemiology 2001, 11, (3), 231–252.

4. Morawska, L.; Allen, J.; Bahnfleth, W.; Bennett, B.; Bluyssen, P. M.; Boerstra, A.; Buonanno, G.; Cao, J. J.; Dancer, S. J.; Floto, A.; Franchimon, F.; Greenhalgh, T.; Haworth, C.; Hogeling, J.; Isaxon, C.; Jimenez, J. L.; Kennedy, A.; Kumar, P.; Kurnitski, J.; Li, Y. G.; Loomans, M.; Marks, G.; Marr, L. C.; Mazzarella, L.; Melikov, A. K.; Miller, S. L.; Milton, D. K.; Monty, J.; Nielsen, P. V.; Noakes, C.; Peccia, J.; Prather, K. A.; Querol, X.; Salthammer, T.; Sekhar, C.; Seppaenen, O.; Tanabe, S.; Tang, J. W.; Tellier, R.; Tham, K. W.; Wargocki, P.; Wierzbicka, A.; Yao, M. S., Mandating indoor air quality for public buildings. Science 2024, 383, (6690), 1418–1420.

5. Morawska, L., The burden of disease due to indoor air pollution and why we need to know about it. Sci Bull 2024, 69, (9), 1161–1164.

6. Liu, N.; Liu, W.; Deng, F.; Liu, Y.; Gao, X.; Fang, L.; Chen, Z.; Tang, H.; Hong, S.; Pan, M.; Liu, W.; Huo, X.; Guo, K.; Ruan, F.; Zhang, W.; Zhao, B.; Mo, J.; Huang, C.; Su, C.; Sun, C.; Zou, Z.; Li, H.; Sun, Y.; Qian, H.; Zheng, X.; Zeng, X.; Guo, J.; Bu, Z.; Mandin, C.; Hanninen, O.; Ji, J. S.; Weschler, L. B.; Kan, H.; Zhao, Z.; Zhang, Y., The burden of disease attributable to indoor air pollutants in China from 2000 to 2017. Lancet Planet Health 2023, 7, (11), e900–e911.

7. Ye, W.; Zhang, X.; Gao, J.; Cao, G. Y.; Zhou, X.; Su, X., Indoor air pollutants, ventilation rate determinants and potential control strategies in Chinese dwellings: A literature review. Sci Total Environ 2017, 586, 696–729.

8. Asikainen, A.; Carrer, P.; Kephalopoulos, S.; Fernandes, E. D.; Wargocki, P.; Hänninen, O., Reducing burden of disease from residential indoor air exposures in Europe (HEALTHVENT project). Environ Health 2016, 15, S35.

9. Liu, Y. M.; Zhou, B.; Wang, J. H.; Zhao, B., Health benefits and cost of using air purifiers to reduce exposure to ambient fine particulate pollution in China. J Hazard Mater 2021, 414, 125540.

10. Yang, K. Q.; Liu, N. R.; Weschler, C. J.; Weschler, L. B.; Mo, J. H.; Xu, Y.; Wei, J. Y.; Wang, Y. M.; Zhao, Z. H.; Kan, H. D.; Zhang, Y. P., Maximizing the net economic benefits of regulating indoor air quality in China. Sustain Cities Soc 2024, 117, 105938.

11. Zhang, A.; Liu, Y. M.; Ji, J. S.; Zhao, B., Air Purifier Intervention to Remove Indoor PM2.5 in Urban China: A Cost-Effectiveness and Health Inequality Impact Study. Environ Sci Technol 2023, 57, (11), 4492–4503.

12. Lewis, A. C.; Jenkins, D.; Whitty, C. J. M., Indoor air pollution: five ways to fight the hidden harms. Nature 2023, 614, (7947), 220–223.

13. Hu, Y.; Ji, J. S.; Zhao, B., Restrictions on indoor and outdoor NO(2) emissions to reduce disease burden for pediatric asthma in China: A modeling study. Lancet Reg Health West Pac 2022, 24, 100463.

14. Hu, Y.; Ji, J. S.; Zhao, B., Deaths Attributable to Indoor PM2.5 in Urban China When Outdoor Air Meets 2021 WHO Air Quality Guidelines. Environ Sci Technol 2022, 56, (22), 15882–15891.

15. Weschler, C. J., Changes in indoor pollutants since the 1950s. Atmos Environ 2009, 43, (1), 153–169.

16. Liu, Z.; Ye, W.; Little, J. C., Predicting emissions of volatile and semivolatile organic compounds from building materials: A review. Build Environ 2013, 64, 7–25.

17. Liu, N. R.; Zhang, X.; Wang, L. Y.; Liang, K.; Zhang, Y. P.; Cao, J. P., Early-Stage Emissions of Formaldehyde and Volatile Organic Compounds from Building Materials: Model Development, Evaluation, and Applications. Environ Sci Technol 2022, 56, (20), 14680–14689.

18. Zhang, Y.; Xiong, J.; Mo, J.; Gong, M.; Cao, J., Understanding and controlling airborne organic compounds in the indoor environment: mass transfer analysis and applications. Indoor Air 2016, 26, (1), 39–60.

19. Zhang, X.; Cao, J. P.; Wei, J. Y.; Zhang, Y. P., Improved C-history method for rapidly and accurately measuring the characteristic parameters of formaldehyde/VOCs emitted from building materials. Build Environ 2018, 143, 570–578.

20. Chen, H. P.; Liu, N. R.; Guo, J.; Wang, L. Y.; Zhang, Y.; Wei, J. Y.; Xu, Y.; Cao, Y. J.; Zhang, Y. P., Two-parameter C-history method: A fast and accurate method for determining the characteristic parameters of formaldehyde/VOC early-stage emissions from building materials. Sci Total Environt 2024, 946, 174218.

21. Huang, S. D.; Xiong, J. Y.; Zhang, Y. P., Impact of Temperature on the Ratio of Initial Emittable Concentration to Total Concentration for Formaldehyde in Building Materials: Theoretical Correlation and Validation. Environ Sci Technol 2015, 49, (3), 1537–1544.

22. Deng, Q. Q.; Yang, X. D.; Zhang, J. S., Study on a new correlation between diffusion coefficient and temperature in porous building materials. Atmos Environ 2009, 43, (12), 2080–2083.

23. Xiong, J. Y.; Zhang, P. P.; Huang, S. D.; Zhang, Y. P., Comprehensive influence of environmental factors on the emission rate of formaldehyde and VOCs in building materials: Correlation development and exposure assessment. Environ Res 2016, 151, 734–741.

24. Hopke, P. K.; Dai, Q. L.; Li, L. X.; Feng, Y. C., Global review of recent source apportionments for airborne particulate matter. Sci Total Environ 2020, 740, 140091.

25. Hopke, P. K.; Feng, Y. C.; Dai, Q. L., Source apportionment of particle number concentrations: A global review. Sci Total Environ 2022, 819, 153104.

26. Saraga, D. E.; Querol, X.; Duarte, R. M. B. O.; Aquilina, N. J.; Canha, N.; Alvarez, E. G.; Jovasevic-Stojanovic, M.; Bekö, G.; Bycenkiene, S.; Kovacevic, R.; Plauskaite, K.; Carslaw, N., Source apportionment for indoor air pollution: Current challenges and future directions. Sci Total Environ 2023, 900, 165744.

27. Minguillón, M. C.; Schembari, A.; Triguero-Mas, M.; de Nazelle, A.; Dadvand, P.; Figueras, F.; Salvado, J. A.; Grimalt, J. O.; Nieuwenhuijsen, M.; Querol, X., Source apportionment of indoor, outdoor and personal PM2.5 exposure of pregnant women in Barcelona, Spain. Atmos Environ 2012, 59, 426–436.

28. Pey, J.; van Drooge, B. L.; Ripoll, A.; Moreno, T.; Grimalt, J. O.; Querol, X.; Alastuey, A., An evaluation of mass, number concentration, chemical composition and types of particles in a cafeteria before and after the passage of an antismoking law. Particuology 2013, 11, (5), 527–532.

29. Barraza, F.; Jorquera, H.; Valdivia, G.; Montoya, L. D., Indoor PM2.5 in Santiago, Chile, spring 2012: Source apportionment and outdoor contributions. Atmos Environ 2014, 94, 692–700.

30. Shang, J.; Khuzestani, R. B.; Tian, J. Y.; Schauer, J. J.; Hua, J. X.; Zhang, Y.; Cai, T. Q.; Fang, D. Q.; An, J. X.; Zhang, Y. X., Chemical characterization and source apportionment of PM2.5 personal exposure of two cohorts living in urban and suburban Beijing. Environ Pollut 2019, 246, 225–236.

31. Tunno, B. J.; Dalton, R.; Cambal, L.; Holguin, F.; Lioy, P.; Clougherty, J. E., Indoor source apportionment in urban communities near industrial sites. Atmos Environ 2016, 139, 30–36.

32. Yang, Y. B.; Liu, L.; Xu, C. Y.; Li, N.; Liu, Z.; Wang, Q.; Xu, D. Q., Source Apportionment and Influencing Factor Analysis of Residential Indoor PM2.5 in Beijing. Int J Env Res Pub He 2018, 15, (4), 686.

33. Zhu, C. S.; Cao, J. J.; Shen, Z. X.; Liu, S. X.; Zhang, T.; Zhao, Z. Z.; Xu, H. M.; Zhang, E. K., Indoor and Outdoor Chemical Components of PM2.5 in the Rural Areas of Northwestern China. Aerosol Air Qual Res 2012, 12, (6), 1157–1165.

34. Brinkman, G. L.; Milford, J. B.; Schauer, J. J.; Shafer, M. M.; Hannigan, M. P., Source identification of personal exposure to fine particulate matter using organic tracers. Atmos Environ 2009, 43, (12), 1972–1981.

35. Carrion-Matta, A.; Kang, C. M.; Gaffin, J. M.; Hauptman, M.; Phipatanakul, W.; Koutrakis, P.; Gold, D. R., Classroom indoor PM2.5 sources and exposures in inner-city schools. Environ Int 2019, 131, 104968.

36. Molnár, P.; Johannesson, S.; Quass, U., Source Apportionment of PM2.5 Using Positive Matrix Factorization (PMF) and PMF with Factor Selection. Aerosol Air Qual Res 2014, 14, (3), 725–733.

37. Amato, F.; Rivas, I.; Viana, M.; Moreno, T.; Bouso, L.; Reche, C.; Alvarez-Pedrerol, M.; Alastuey, A.; Sunyer, J.; Querol, X., Sources of indoor and outdoor PM2.5 concentrations in primary schools. Sci Total Environ 2014, 490, 757–765.

38. Bari, M. A.; Kindzierski, W. B.; Wheeler, A. J.; Héroux, M. E.; Wallace, L. A., Source apportionment of indoor and outdoor volatile organic compounds at homes in Edmonton, Canada. Build Environ 2015, 90, 114–124.

39. Huang, Y.; Su, T.; Wang, L. Q.; Wang, N.; Xue, Y. G.; Dai, W. T.; Lee, S. C.; Cao, J. J.; Ho, S. S. H., Evaluation and characterization of volatile air toxics indoors in a heavy polluted city of northwestern China in wintertime. Sci Total Environ 2019, 662, 470–480.

40. Huang, L. H.; Qian, H.; Deng, S. X.; Guo, J. F.; Li, Y. P.; Zhao, W. P.; Yue, Y., Urban residential indoor volatile organic compounds in summer, Beijing: Profile, concentration and source characterization. Atmos Environ 2018, 188, 1–11.

41. Liu, Q. Y.; Liu, Y. J.; Zhang, M. G., Source Apportionment of Personal Exposure to Carbonyl Compounds and BTEX at Homes in Beijing, China. Aerosol Air Qual Res 2014, 14, (1), 330–337.

42. Norris, C.; Fang, L.; Barkjohn, K. K.; Carlson, D.; Zhang, Y. P.; Mo, J. H.; Li, Z.; Zhang, J. F.; Cui, X. X.; Schauer, J. J.; Davis, A.; Black, M.; Bergin, M. H., Sources of volatile organic compounds in suburban homes in Shanghai, China, and the impact of air filtration on compound concentrations. Chemosphere 2019, 231, 256–268.

43. Rösch, C.; Kohajda, T.; Röder, S.; von Bergen, M.; Schlink, U., Relationship between sources and patterns of VOCs in indoor air. Atmos Pollut Res 2014, 5, (1), 129–137.

44. Guo, H., Source apportionment of volatile organic compounds in Hong Kong homes. Build Environ 2011, 46, (11), 2280–2286.

45. Jia, C.; Batterman, S.; Godwin, C., VOCs in industrial, urban and suburban neighborhoods - Part 2: Factors affecting indoor and outdoor concentrations. Atmos Environ 2008, 42, (9), 2101–2116.

46. Lunderberg, D. M.; Misztal, P. K.; Liu, Y. J.; Arata, C.; Tian, Y. L.; Kristensen, K.; Weber, R. J.; Nazaroff, W. W.; Goldstein, A. H., High-Resolution Exposure Assessment for Volatile Organic Compounds in Two California Residences. Environ Sci Technol 2021, 55, (10), 6740–6751.

47. Dai, X. L.; Liu, J. J.; Yin, Y. H.; Song, X.; Jia, S. S., Modeling and controlling indoor formaldehyde concentrations in apartments: On-site investigation in all climate zones of China. Build Environ 2018, 127, 98–106.

48. Pei, J. J.; Yin, Y. H.; Liu, J. J.; Dai, X. L., An eight-city study of volatile organic compounds in Chinese residences: Compounds, concentrations, and characteristics. Sci Total Environ 2020, 698, 134137.

49. U.S. Environmental Protection Agency EPA Positive Matrix Factorization (PMF) 5.0 Fundamentals and User Guide; 2014.

50. Liu, N.; Oshan, R.; Blanco, M.; Sheppard, L.; Seto, E.; Larson, T.; Austin, E., Mapping Source-Specific Air Pollution Exposures Using Positive Matrix Factorization Applied to Multipollutant Mobile Monitoring in Seattle, WA. Environ Sci Technol 2025, 59, (7), 3443–3458.

51. Lee, E.; Chan, C. K.; Paatero, P., Application of positive matrix factorization in source apportionment of particulate pollutants in Hong Kong. Atmos Environ 1999, 33, (19), 3201–3212.

52. Baudic, A.; Gros, V.; Sauvage, S.; Locoge, N.; Sanchez, O.; Sarda-Estève, R.; Kalogridis, C.; Petit, J. E.; Bonnaire, N.; Baisnée, D.; Favez, O.; Albinet, A.; Sciare, J.; Bonsang, B., Seasonal variability and source apportionment of volatile organic compounds (VOCs) in the Paris megacity (France). Atmos Chem Phys 2016, 16, (18), 11961–11989.

53. Liao, H. T.; Yau, Y. C.; Huang, C. S.; Chen, N.; Chow, J. C.; Watson, J. G.; Tsai, S. W.; Chou, C. C. K.; Wu, C. F., Source apportionment of urban air pollutants using constrained receptor models with a priori profile information. Environ Pollut 2017, 227, 323–333.

54. Wang, M.; Wang, Q. Y.; Ho, S. S. H.; Li, H.; Zhang, R. J.; Ran, W. K.; Qu, L. L.; Lee, S. C.; Cao, J. J., Chemical characteristics and sources of nitrogen-containing organic compounds at a regional site in the North China Plain during the transition period of autumn and winter. Sci Total Environ 2022, 812, 151451.

55. Huang, C. S.; Liu, Y. H.; Liao, H. T.; Chen, C. Y.; Wu, C. F., Improvements in source apportionment of multiple time-resolved PM(2.5) inorganic and organic speciation measurements using constrained Positive Matrix Factorization. Environ Sci Pollut Res Int 2024, 31, (55), 64185–64198.

56. Huang, C. S.; Liao, H. T.; Lu, S. H.; Chan, C. C.; Wu, C. F., Identifying and quantifying PM(2.5) pollution episodes with a fusion method of moving window technique and constrained Positive Matrix Factorization. Environ Pollut 2022, 315, 120382.

57. Brown, S. G.; Eberly, S.; Paatero, P.; Norris, G. A., Methods for estimating uncertainty in PMF solutions: Examples with ambient air and water quality data and guidance on reporting PMF results. Sci Total Environ 2015, 518, 626–635.

58. Paatero, P.; Eberly, S.; Brown, S. G.; Norris, G. A., Methods for estimating uncertainty in factor analytic solutions. Atmos Meas Tech 2014, 7, (3), 781–797.

59. Wu, C. F.; Wu, S. Y.; Wu, Y. H.; Cullen, A. C.; Larson, T. V.; Williamson, J.; Liu, L. J. S., Cancer risk assessment of selected hazardous air pollutants in Seattle. Environ Int 2009, 35, (3), 516–522.

60. Liao, H. T.; Chou, C. C. K.; Chow, J. C.; Watson, J. G.; Hopke, P. K.; Wu, C. F., Source and risk apportionment of selected VOCs and PM2.5 species using partially constrained receptor models with multiple time resolution data. Environ Pollut 2015, 205, 121–130.

61. Liu, N. R.; Bu, Z. M.; Liu, W.; Kan, H. D.; Zhao, Z. H.; Deng, F. R.; Huang, C.; Zhao, B.; Zeng, X. G.; Sun, Y. X.; Qian, H.; Mo, J. H.; Sun, C. J.; Guo, J. G.; Zheng, X. H.; Weschler, L. B.; Zhang, Y. P., Health effects of exposure to indoor volatile organic compounds from 1980 to 2017: A systematic review and meta-analysis. Indoor Air 2022, 32, (5), e13038.

62. Liu, N. R.; Fang, L.; Liu, W.; Kan, H. D.; Zhao, Z. H.; Deng, F. R.; Huang, C.; Zhao, B.; Zeng, X. A.; Sun, Y. X.; Qian, H.; Mo, J. H.; Sun, C. J.; Guo, J. G.; Zheng, X. H.; Bu, Z. M.; Weschler, L. B.; Zhang, Y. P., Health effects of exposure to indoor formaldehyde in civil buildings: A systematic review and meta-analysis on the literature in the past 40 years. Build Environ 2023, 233, 110080.

63. California Environmental Protection Agency Toxicity criteria on chemicals evaluated by Office of Environmental Health Hazard Assessment (OEHHA). https://oehha.ca.gov/library/chemicals (Apr 8, 2025),

64. Department of Environment, G. L., and Energy in Michigan,, Air Toxics Program. https://www.michigan.gov/egle/about/organization/air-quality/air-toxics (Apr 8, 2025),

65. U.S. Environmental Protection Agency Integrated Risk Information System (IRIS) Assessments. https://iris.epa.gov/AtoZ/?list_type=alpha (Apr 8, 2025),

66. Hänninen, O.; Knol, A. B.; Jantunen, M.; Lim, T. A.; Conrad, A.; Rappolder, M.; Carrer, P.; Fanetti, A. C.; Kim, R.; Buekers, J.; Torfs, R.; Iavarone, I.; Classen, T.; Hornberg, C.; Mekel, O. C. L.; Grp, E. W., Environmental Burden of Disease in Europe: Assessing Nine Risk Factors in Six Countries. Environ Health Persp 2014, 122, (5), 439–446.

67. Xiong, Y.; Du, K.; Huang, Y. X., One-third of global population at cancer risk due to elevated volatile organic compounds levels. Npj Clim Atmos Sci 2024, 7, (1), 54.

68. Greco, S. L.; MacIntyre, E.; Young, S.; Warden, H.; Drudge, C.; Kim, J.; Candido, E.; Demers, P.; Copes, R., An approach to estimating the environmental burden of cancer from known and probable carcinogens: application to Ontario, Canada. BMC Public Health 2020, 20, (1), 1017.

69. Zhou, M. G.; Wang, H. D.; Zeng, X. Y.; Yin, P.; Zhu, J.; Chen, W. Q.; Li, X. H.; Wang, L. J.; Wang, L. M.; Liu, Y. N.; Liu, J. M.; Zhang, M.; Qi, J. L.; Yu, S. C.; Afshin, A.; Gakidou, E.; Glenn, S.; Krish, V. S.; Miller-Petrie, M. K.; Mountjoy-Venning, W. C.; Mullany, E. C.; Redford, S. B.; Liu, H. Y.; Naghavi, M.; Hay, S. I.; Wang, L. H.; Murray, C. J. L.; Liang, X. F., Mortality, morbidity, and risk factors in China and its provinces, 1990-2017: a systematic analysis for the Global Burden of Disease Study 2017. Lancet 2019, 394, (10204), 1145–1158.

70. Bellis, M. A.; Hughes, K.; Ford, K.; Rodriguez, G. R.; Sethi, D.; Passmore, J., Life course health consequences and associated annual costs of adverse childhood experiences across Europe and North America: a systematic review and meta-analysis. Lancet Public Health 2019, 4, (10), E517–E528.

71. Hughes, K.; Ford, K.; Bells, M. A.; Glendinning, F.; Harrison, E.; Passmore, J., Health and financial costs of adverse childhood experiences in 28 European countries: a systematic review and meta-analysis. Lancet Public Health 2021, 6, (11), E848–E857.

72. Alapieti, T.; Mikkola, R.; Pasanen, P.; Salonen, H., The influence of wooden interior materials on indoor environment: a review. Eur J Wood Prod 2020, 78, (4), 617–634.

73. Böhm, M.; Salem, M. Z. M.; Srba, J., Formaldehyde emission monitoring from a variety of solid wood, plywood, blockboard and flooring products manufactured for building and furnishing materials. J Hazard Mater 2012, 221, 68–79.

74. Harb, P.; Locoge, N.; Thevenet, F., Emissions and treatment of VOCs emitted from wood-based construction materials: Impact on indoor air quality. Chem Eng J 2018, 354, 641–652.

75. Jiang, C. J.; Li, D. D.; Zhang, P. Y.; Li, J. G.; Wang, J.; Yu, J. G., Formaldehyde and volatile organic compound (VOC) emissions from particleboard: Identification of odorous compounds and effects of heat treatment. Build Environ 2017, 117, 118–126.

76. Wang, Y. Z.; Wang, H. M.; Tan, Y. D.; Liu, J. L.; Wang, K. L.; Ji, W. J.; Sun, L. H.; Yu, X. F.; Zhao, J.; Xu, B. P.; Xiong, J. Y., Measurement of the key parameters of VOC emissions from wooden furniture, and the impact of temperature. Atmos Environ 2021, 259, 118510.

77. Liang, W. H.; Lv, M. Q.; Yang, X. D., The effect of humidity on formaldehyde emission parameters of a medium-density fiberboard: Experimental observations and correlations. Build Environ 2016, 101, 110–115.

78. Fang, L.; Liu, N. R.; Liu, W.; Mo, J. H.; Zhao, Z. H.; Kan, H. D.; Deng, F. R.; Huang, C.; Zhao, B.; Zeng, X. G.; Sun, Y. X.; Qian, H.; Sun, C. J.; Guo, J. G.; Zheng, X. H.; Zhang, Y. P., Indoor formaldehyde levels in residences, schools, and offices in China in the past 30 years: A systematic review. Indoor Air 2022, 32, (10), e13141.

79. Yuan, B.; Shao, M.; Lu, S. H.; Wang, B., Source profiles of volatile organic compounds associated with solvent use in Beijing, China. Atmos Environ 2010, 44, (15), 1919–1926.

80. Pei, Y. P.; Liu, N.; Liu, S. H.; Guan, H. Y.; Guo, Z. B.; Li, Q. N.; Han, W.; Cai, H. M., Investigation of odor emissions from coating products: Key factors and key odorants. Front Env Sci 2022, 10, 1039842.

81. Qi, Y. Q.; Shen, L. M.; Zhang, J. L.; Yao, J.; Lu, R.; Miyakoshi, T., Species and release characteristics of VOCs in furniture coating process. Environ Pollut 2019, 245, 810–819.

82. Tong, R. P.; Zhang, L.; Yang, X. Y.; Liu, J. F.; Zhou, P. N.; Li, J. F., Emission characteristics and probabilistic health risk of volatile organic compounds from solvents in wooden furniture manufacturing. J Clean Prod 2019, 208, 1096–1108.

83. Cox, S. S.; Little, J. C.; Hodgson, A. T., Measuring concentrations of volatile organic compounds in vinyl flooring. J Air Waste Manag Assoc 2001, 51, (8), 1195–1201.

84. Cox, S. S.; Little, J. C.; Hodgson, A. T., Predicting the emission rate of volatile organic compounds from vinyl flooring. Environ Sci Technol 2002, 36, (4), 709–714.

85. Kraus, M.; Senitkova, I. J., VOCs Emission Simulation of Common Flooring Materials. IOP Conf Ser Mater Sci Eng 2020, 960, (4), 042093.

86. Wilke, O.; Jann, O.; Brodner, D., VOC- and SVOC-emissions from adhesives, floor coverings and complete floor structures. Indoor Air 2004, 14 *Suppl 8*, 98–107.

87. He, G.; Yang, X. D.; Shaw, C. Y., Material emission parameters obtained through regression. Indoor Built Environ 2005, 14, (1), 59–68.

88. Wakayama, T.; Ito, Y.; Sakai, K.; Miyake, M.; Shibata, E.; Ohno, H.; Kamijima, M., Comprehensive review of 2-ethyl-1-hexanol as an indoor air pollutant. J Occup Health 2019, 61, (1), 19–35.

89. Milhem, S. A.; Verriele, M.; Nicolas, M.; Thevenet, F., Does the ubiquitous use of essential oil-based products promote indoor air quality? A critical literature review. Environ Sci Pollut R 2020, 27, (13), 14365–14411.

90. Thevenet, F.; Verriele, M.; Harb, P.; Thlaijeh, S.; Brun, R.; Nicolas, M.; Angulo-Milhem, S., The indoor fate of terpenes: Quantification of the limonene uptake by materials. Build Environ 2021, 188, 107433.

91. Kim, Y. W.; Kim, M. J.; Chung, B. Y.; Bang, D. Y.; Lim, S. K.; Choi, S. M.; Lim, D. S.; Cho, M. C.; Yoon, K.; Kim, H. S.; Kim, K. B.; Kim, Y. S.; Kwack, S. J.; Lee, B. M., Safety Evaluation And Risk Assessment Of d-Limonene. J Toxicol Env Heal B 2013, 16, (1), 17–38.

92. Coggon, M. M.; Stockwell, C. E.; Xu, L.; Peischl, J.; Gilman, J. B.; Lamplugh, A.; Bowman, H. J.; Aikin, K.; Harkins, C.; Zhu, Q. D.; Schwantes, R. H.; He, J.; Li, M.; Seltzer, K.; McDonald, B.; Warneke, C., Contribution of cooking emissions to the urban volatile organic compounds in Las Vegas, NV. Atmos Chem Phys 2024, 24, (7), 4289–4304.

93. Katragadda, H. R.; Fullana, A.; Sidhu, S.; Carbonell-Barrachina, A. A., Emissions of volatile aldehydes from heated cooking oils. Food Chem 2010, 120, (1), 59–65.

94. Li, L. X.; Cheng, Y.; Dai, Q. L.; Liu, B. S.; Wu, J. H.; Bi, X. H.; Choe, T. H.; Feng, Y. C., Chemical characterization and health risk assessment of VOCs and PM2.5-bound PAHs emitted from typical Chinese residential cooking. Atmos Environ 2022, 291, 119392.

95. Zhang, D. C.; Liu, J. J.; Jia, L. Z.; Wang, P.; Han, X., Speciation of VOCs in the cooking fumes from five edible oils and their corresponding health risk assessments. Atmos Environ 2019, 211, 6–17.

96. Zhang, W.; Bai, Z.; Shi, L. B.; Son, J. H.; Li, L.; Wang, L. N.; Chen, J. M., Investigating aldehyde and ketone compounds produced from indoor cooking emissions and assessing their health risk to human beings. J Environ Sci 2023, 127, 389–398.

97. Peng, C. Y.; Lang, C. H.; Lin, P. C.; Kuo, Y. C., Effects of cooking method, cooking oil, and food type on aldehyde emissions in cooking oil fumes. J Hazard Mater 2017, 324, 160–167.

98. Deng, H. F.; Qiu, J.; Zhang, R. Q.; Xu, J. L.; Qu, Y. K.; Wang, J. X.; Liu, Y. J.; Gligorovski, S., Ozone Chemistry on Greasy Glass Surfaces Affects the Levels of Volatile Organic Compounds in Indoor Environments. Environ Sci Technol 2024, 58, (19), 8393–8403.

99. Wang, H.; Morrison, G., Ozone-surface reactions in five homes: surface reaction probabilities, aldehyde yields, and trends. Indoor Air 2010, 20, (3), 224–234.

100. Wang, H.; Morrison, G. C., Ozone-initiated secondary emission rates of aldehydes from indoor surfaces in four homes. Environ Sci Technol 2006, 40, (17), 5263–5268.

101. Song, K.; Tang, R. Z.; Li, A.; Wan, Z. C.; Zhang, Y.; Gong, Y. Z.; Lv, D. Q.; Lu, S. H.; Tan, Y.; Yan, S. Y.; Yan, S. C.; Zhang, J. S.; Fan, B. M.; Chan, C. K.; Guo, S., Particulate organic emissions from incense-burning smoke: Chemical compositions and emission characteristics. Sci Total Environ 2023, 897, 165319.

102. Manoukian, A.; Quivet, E.; Temime-Roussel, B.; Nicolas, M.; Maupetit, F.; Wortham, H., Emission characteristics of air pollutants from incense and candle burning in indoor atmospheres. Environ Sci Pollut R 2013, 20, (7), 4659–4670.

103. Pang, X. B.; Lewis, A. C., Carbonyl compounds in gas and particle phases of mainstream cigarette smoke. Sci Total Environ 2011, 409, (23), 5000–5009.

104. Baghani, A. N.; Dana, E.; Sorooshian, A.; Jafari, A. J.; Aalamolhoda, A. A.; Sheikhi, R.; Jajarmi, F.; Shahsavani, A.; Delikhoon, M.; Ebrahimzade, G.; Ashournejad, Q.; Mansoorian, H. J.; Kermani, M., Sensitivity of BTEX pollution and health effects to traffic restrictions: A case study in an urban center of Tehran, Iran. Sustain Cities Soc 2024, 104, 105281.

105. Hun, D. E.; Corsi, R. L.; Morandi, M. T.; Siegel, J. A., Automobile proximity and indoor residential concentrations of BTEX and MTBE. Build Environ 2011, 46, (1), 45–53.

106. Li, B. W.; Wang, J. N.; Wang, J. L.; Zhang, L. J.; Zhang, Q. Y., A comprehensive study on emission of volatile organic compounds for light duty gasoline passenger vehicles in China: Illustration of impact factors and renewal emissions of major compounds. Environ Res 2021, 193, 110461.

107. Truc, V. T. Q.; Oanh, N. T. K., Roadside BTEX and other gaseous air pollutants in relation to emission sources. Atmos Environ 2007, 41, (36), 7685–7697.

108. Ceron-Breton, J. G.; Bretón, R. M. C.; Kahl, J. D. W.; Rico, G. S.; Lozada, S. E. C.; Fuentes, M. D. E.; Chi, M. P. U., Concentrations, sources, and health risk associated with exposure to BTEX at ten sites located in an urban-industrial area in the Bajio Region, Mexico. Air Qual Atmos Hlth 2021, 14, (5), 741–761.

109. Horton, D. E.; Harshvardhan; Diffenbaugh, N. S., Response of air stagnation frequency to anthropogenically enhanced radiative forcing. Environ Res Lett 2012, 7, (4), 044034.

110. Khoshakhlagh, A. H.; Yazdanirad, S.; Sicard, P., Exposure to BTEX concentrations in different indoor microenvironments: Emphasis on different times of the year. Risk Anal 2025, 70032.

111. Zhang, Z.; Man, H.; Zhao, J.; Jiang, Y.; Zeng, M.; Cai, Z.; Huang, C.; Huang, W.; Zhao, H.; Jing, S.; Shi, X.; He, K.; Liu, H., Primary organic gas emissions in vehicle cold start events: Rates, compositions and temperature effects. J Hazard Mater 2022, 435, 128979.

112. Liu, Y.; Shao, M.; Fu, L. L.; Lu, S. H.; Zeng, L. M.; Tang, D. G., Source profiles of volatile organic compounds (VOCs) measured in China: Part I. Atmos Environ 2008, 42, (25), 6247–6260.

113. Jiang, C. Y.; Pei, C. L.; Cheng, C. L.; Shen, H. Z.; Zhang, Q. H.; Lian, X. F.; Xiong, X.; Gao, W.; Liu, M.; Wang, Z. X.; Huang, B.; Tang, M.; Yang, F.; Zhou, Z.; Li, M., Emission factors and source profiles of volatile organic compounds from typical industrial sources in Guangzhou, China. Sci Total Environ 2023, 869, 161758.

114. Meng, L. N.; Gao, S.; Zhang, S. W.; Che, X.; Jiao, Z.; Ren, Y.; Wang, C. G., Identification of atmospheric emerging contaminants from industrial emissions: A case study of halogenated hydrocarbons emitted by the pharmaceutical industry. Environ Int 2024, 192, 109027.

115. Cheng, N. N.; Jing, D. J.; Zhang, C.; Chen, Z. W.; Li, W.; Li, S. J.; Wang, Q. L., Process-based VOCs source profiles and contributions to ozone formation and carcinogenic risk in a typical chemical synthesis pharmaceutical industry in China. Sci Total Environ 2021, 752, 141899.

116. Cui, L. L.; Wu, D.; Wang, S. X.; Xu, Q. C.; Hu, R. L.; Hao, J. M., Measurement report: Ambient volatile organic compound (VOC) pollution in urban Beijing: characteristics, sources, and implications for pollution control. Atmos Chem Phys 2022, 22, (18), 11931–11944.

117. Mo, Z. W.; Shao, M.; Lu, S. H., Compilation of a source profile database for hydrocarbon and OVOC emissions in China. Atmos Environ 2016, 143, 209–217.

118. Zhang, Y. L.; Wang, X. M.; Zhang, Z.; Lü, S. J.; Shao, M.; Lee, F. S. C.; Yu, J. Z., Species profiles and normalized reactivity of volatile organic compounds from gasoline evaporation in China. Atmos Environ 2013, 79, 110–118.

119. Pekey, H.; Pekey, B.; Arslanbaş, D.; Bozkurt, Z. B.; Doğan, G.; Tuncel, G., Source Apportionment of Personal Exposure to Fine Particulate Matter and Volatile Organic Compounds using Positive Matrix Factorization. Water Air Soil Poll 2012, 224, (1), 1403.

120. Daisey, J. M., Tracers for assessing exposure to environmental tobacco smoke: What are they tracing? Environ Health Persp 1999, 107, 319–327.

121. Simoneit, B. R. T., Biomass burning - A review of organic tracers for smoke from incomplete combustion. Appl Geochem 2002, 17, (3), 129–162.

122. Liu, Z. H.; Su, J. B.; Ma, A. J.; Zhu, A. X.; Liu, P. Y., Study on emission characteristics of tracer pollutants in cooking oil fumes. Atmos Pollut Res 2022, 13, (5), 101409.

123. Huang, D. D.; Zhu, S. H.; An, J. Y.; Wang, Q. Q.; Qiao, L. P.; Zhou, M.; He, X.; Ma, Y. G.; Sun, Y. L.; Huang, C.; Yu, J. Z.; Zhang, Q., Comparative Assessment of Cooking Emission Contributions to Urban Organic Aerosol Using Online Molecular Tracers and Aerosol Mass Spectrometry Measurements. Environ Sci Technol 2021, 55, (21), 14526–14535.

