## Supplementary Material 1 for "Source-specific exposure and burden of disease attributable to volatile organic compounds (VOCs) in China’s residences"

This supporting information includes 6 sections, 13 figures, and 7 tables.

Tables S8-S20 are available in **Supporting Information 2**.

### Table of contents


**Section S1 Data collection and pre-processing .................................... S3**

S1.1 Data collection ........................................................................................ S3
S1.2 Missing data imputation in data pre-processing ..................................... S4

**Section S2 PMF analysis methods ........................................................ S6**

S2.1 PMF modeling ........................................................................................ S6
S2.2 PMF diagnosis ........................................................................................ S8

**Section S3 Burden of disease estimation methods ............................. S11**

S3.1 Exposure-response (E-R) relationships ................................................ S11
S3.2 Total DALYs, baseline incidence and other data .................................. S13
S3.3 Uncertainty analysis ............................................................................. S14

**Section S4 Source-specific exposure assessment ............................... S16**

S4.1 Source apportionment ........................................................................... S16
S4.2 Source-specific contribution to VOC concentrations ............................ S20
S4.3 Multi-room VOC concentration comparison ......................................... S22
S4.4 PMF model performance for different cities ........................................ S24

**Section S5 Source-specific burden of disease .................................... S25**

**Section S6 Comparison with previous studies ................................... S29**

**Reference ............................................................................................ S30**

### Section S1 Data collection and pre-processing

#### S1.1 Data collection

**Figure S1.** Location of 9 included cities for residential VOC measurements in China.

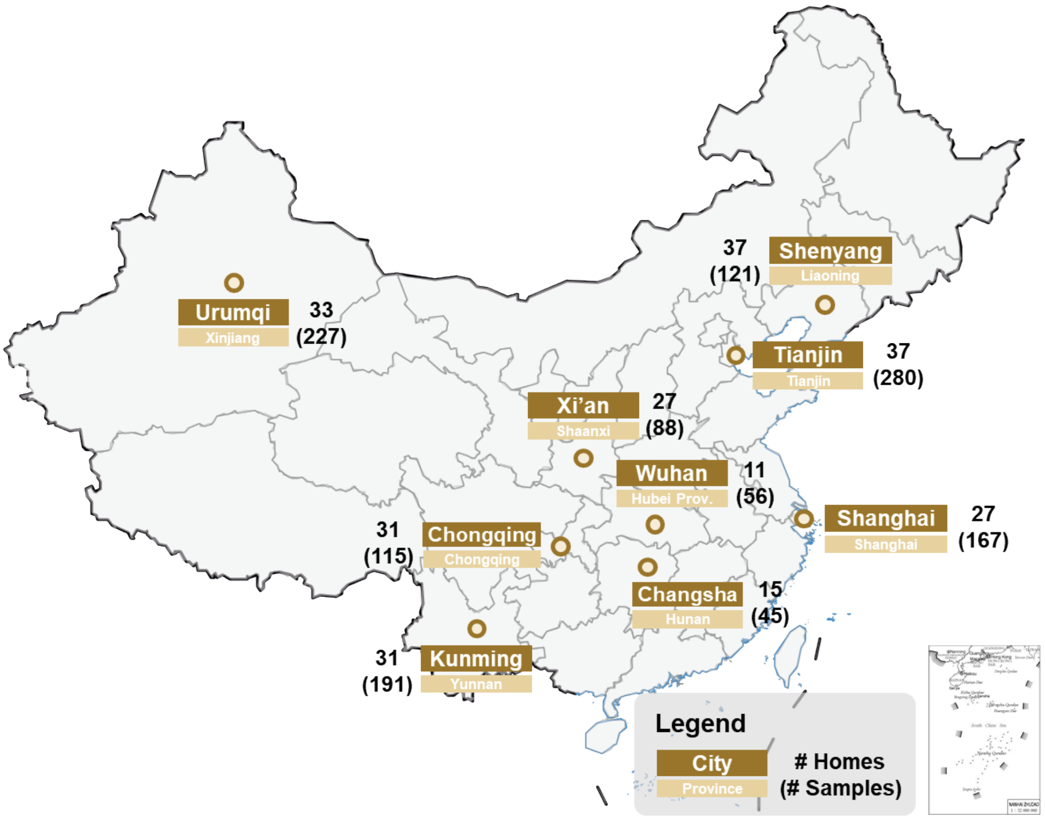


**Table S1.** VOCs included in the PMF analysis.

| Type | Pollutant | CAS | Detection ratio |
|---|---|---|---|
| Alkane | Isobutane | 75-28-5 | 32.2% |
| | Pentane | 109-66-0 | 28.6% |
| | Isopentane | 78-78-4 | 74.7% |
| | n-Hexane | 110-54-3 | 74.3% |
| | Heptane | 142-82-5 | 36.4% |
| | Undecane | 1120-21-4 | 34.5% |
| | Tridecane | 629-50-5 | 31.1% |
| | Tetradecane | 629-59-4 | 50.5% |
| Halogenated organic compounds | Dichloromethane (DCM) | 75-09-2 | 50.7% |
| | 1,2-Dichloroethane (DCE) | 107-06-2 | 39.8% |
| Alcohol | Ethanol | 64-17-5 | 46.6% |
| | 1-Butanol | 71-36-3 | 68.8% |
| | 2-Ethyl-1-hexanol | 104-76-7 | 62.4% |
| Aldehyde & Ketone | Formaldehyde | 50-00-0 | 100.0% |

| Type | Pollutant | CAS | Detection ratio |
|---|---|---|---|
| | Acetaldehyde | 75-07-0 | 50.7% |
| | Pentanal | 110-62-3 | 53.8% |
| | Hexanal | 66-25-1 | 88.8% |
| | Heptanal | 111-71-7 | 50.5% |
| | Octanal | 124-13-0 | 46.5% |
| | Nonanal | 124-19-6 | 83.5% |
| | Decanal | 112-31-2 | 64.3% |
| | Acetone | 67-64-1 | 40.5% |
| | 2-Butanone | 78-93-3 | 54.5% |
| | Cyclohexanone | 108-94-1 | 51.0% |
| Acetate | Ethyl acetate | 141-78-6 | 77.5% |
| | Butyl acetate | 123-86-4 | 76.7% |
| | sec-Butyl acetate | 105-46-4 | 46.0% |
| | n-Butyl acrylate | 141-32-2 | 34.5% |
| | 1-Methoxy-2-propyl acetate (PGMEA) | 108-65-6 | 32.7% |
| Aromatic hydrocarbon | Benzene | 71-43-2 | 86.0% |
| | Toluene | 108-88-3 | 90.1% |
| | Ethylbenzene | 100-41-4 | 71.4% |
| | Xylenes | 108-38-3<br>95-47-6<br>106-42-3 | 95.0% |
| | Styrene | 100-42-5 | 33.6% |
| | Benzaldehyde | 100-52-7 | 69.5% |
| | Naphthalene | 91-20-3 | 40.9% |
| Terpene | Limonene | 138-86-3 | 45.1% |
| | α-Pinene | 80-56-8 | 30.6% |
| Carboxylic acid | Acetic acid | 64-19-7 | 31.6% |

#### S1.2 Missing data imputation in data pre-processing

There are two different types of missing data in this study. The first type happened in the samples in winter and spring in Shanghai, Changsha, and Wuhan. For these samples, only 9 criteria VOCs were measured, including formaldehyde, benzene, toluene, ethylbenzene, xylenes, styrene, undecane, butyl acetate, and TVOCs. Other VOCs were not analyzed. If the concentrations in the same residence in other seasons were available, we used their medians to impute the input concentration matrix (Scenario 1A). Otherwise, we used medians of all residences in other seasons in that city for the imputation (Scenario 1B). The imputed uncertainty was 4 times that the imputed concentration. [1]

The second type of missing data was about the measured formaldehyde concentrations. Since formaldehyde and VOC measurements were obtained from different methods and teams, the two datasets sometimes cannot match with each other, which means that the a few formaldehyde concentrations were missing for some samples. If the formaldehyde concentrations in the same residence and same season but different rooms were available, the median of these concentrations were used for imputation (Scenario 2A). Otherwise, we used median concentration of all residences in that city in the same season for imputation (Scenario 2B). When providing the uncertainty for these missing data, the uncertainty in scenario 2B was imputed by 4 times that the imputed concentration, because we consider them as completely missing. [1] However, we used the following equation for scenario 2A.

$$Unc = \left[1.1\times\left(1+3.55\min\left\{RD,100\%\right\}\right)-1\right]\times Conc \tag{S1}$$

where *RD* is the median relative difference in formaldehyde concentration between the room in the focused sample and other room types. We designed this formula because we think that the formaldehyde concentrations in bedrooms, kitchens, and living rooms were usually similar/correlated in the same residence and the same season due to the air circulation indoors. Therefore, we did not consider this scenario as a completely missing scenario like Scenario 1A, 1B, and 2B. The value 1.1 in Equation (S1) refers to 10% error fraction used in the uncertainty estimation in Equation (4) in the main text. The relative deviation between different room types was considered as an additional contribution to that error fraction. When the relative deviation is large enough to exceed 100%, we considered it as completely missing. Therefore, when the *RD* equals 0, it suggests formaldehyde imputed concentrations are completely accurate, so Unc = 0.1Conc, which is approximately as Equation (4) in the main text. When the RD equals to 100%, it suggests the data are completely missing, so Unc = 4Conc, which is the same as the uncertainty imputation formula for scenario 1A, 1B, and 2B. That's why we selected 3.55 in Equation (S1).

### Section S2 PMF analysis methods

#### S2.1 PMF modeling

We performed all PMF analysis using the EPA PMF 5.0 software in a spatiotemporal context. All 1290 samples from 249 residences in nine cities across four different seasons were included in one PMF model (i.e., $I$ = 1290). In addition to 39 VOCs, TVOC concentrations were also included, which was the sum of formaldehyde concentrations and all detected VOC concentrations in the Tenax-TA tubes (i.e., $J$ = 40). Following EPA PMF guidance, each VOC species was categorized as ‘strong’, ‘weak’, or ‘bad’ based on signal-to-noise (S/N) ratio. [1, 2] Species with S/N < 0.5 were excluded (“bad”), those with 0.5 ≤ S/N < 1 were designated “weak” (with tripled uncertainties), and species with S/N ≥ 1 were considered “strong”. In this study, all 39 included VOCs were “strong” species. TVOCs were categorized as “weak” as it was regarded as the total variable.

In this study, we conducted the PMF modeling through the following steps:

1) We tried running PMF models with 3 to 10 factors separately, and calculated the maximum individual column mean (IM) and standard deviation (IS) of the scaled residuals.
2) We selected the number of factors (i.e., $K$ in Equation (1) in the main text) where IM and IS had a significant drop as the number of factors increase. [3-6]
3) With the selected number of factors, we calculated the Pearson and Spearman correlation coefficient between the observed and the predicted concentrations for each pollutant. If both correlation coefficients were lower than 0.6, it suggests a poor model fit for this pollutant, which was then downweighed from “strong” to “weak” species. [7, 8] It should be noted that the VOCs with health risk data and a detection ratio over 60% were considered as priority pollutants and kept strong

(including formaldehyde, benzene, toluene, ethylbenzene, and xylenes).

4) We finally checked the interpretability of factor profiles and bootstrap results (also seen in **Section S2.2**). If they did not satisfy the corresponding requirements of bootstrap, we would then try ($K$-1) or ($K$+1) factors and repeat steps 1) – 4). If the interpretability of some factors was not good enough, we considered additional constraints for factor rotation.

According to **Figure S2** below, the IM and IS had a significant drop when the number of factors was equal to 9. Therefore, we initially selected the PMF model with 9 factors. After calculating the correlations between observed and predicted concentrations, 12 VOCs were downweighed from “strong” to “weak” species, including isobutane, heptane, undecane, dichloromethane, ethanol, acetaldehyde, acetone, n-butyl acrylate, styrene, naphthalene, α-pinene, and acetic acid. After checking the interpretability of derived factor profiles, we decided to pull up the formaldehyde and benzene concentrations (originally zero in the base profile) maximally but constrained d$Q$% within 0.5% in the profile of cooking and indoor combustion source. That’s because previous studies show that cooking and indoor combustion can emit considerable amount of formaldehyde and benzene into indoor air.[9-12] The solution after this rotation was called as the constrained solution.

**Figure S2.** IM and IS for PMF models with different numbers of factors.

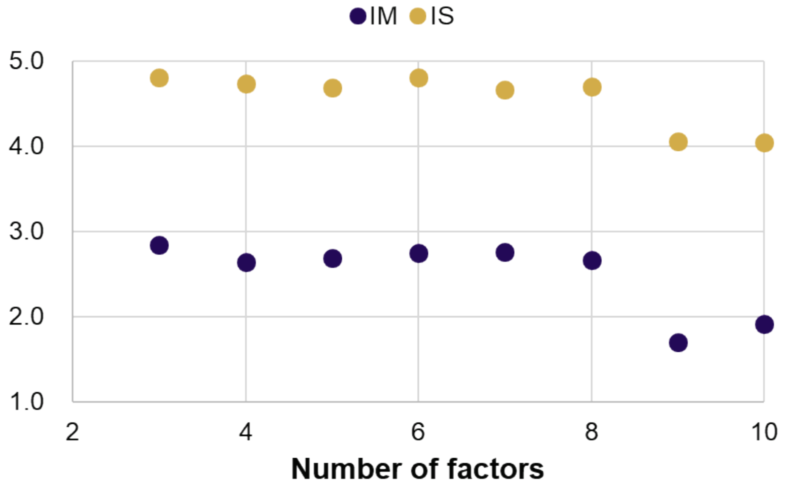

#### S2.2 PMF diagnosis

**1) Block bootstrap**

We generated 100 block bootstrap samples according to the sample size in different cities and ran PMF models separately. It means that, for each bootstrap sample, we included 280, 121, 167, 45, 56, 115, 191, 88, and 227 samples for Tianjin, Shenyang, Shanghai, Changsha, Wuhan, Chongqing, Kunming, Xi'an, and Urumqi, respectively. Then each block bootstrap sample was input into EPA PMF 5.0 software manually to derive the factor profile and contribution matrices. As is shown in **Table S2** below, the accuracy of mapping bootstrap factors to base factors was over 80% for all 9 factors. Among them, 5 factors had a 100% mapping accuracy, while factor 5 (fragranced products) had the lowest mapping accuracy (82%). The bootstrap results suggest that the PMF solutions are robust enough, and the assumption of spatiotemporal constant profiles was appropriate.

**Table S2.** Results of block bootstrap for the constrained solution.

| | Fac1 | Fac2 | Fac3 | Fac4 | Fac5 | Fac6 | Fac7 | Fac8 | Fac9 | Unmapped | Accuracy |
|---|---|---|---|---|---|---|---|---|---|---|---|
| Boot Fac1 | 100 | 0 | 0 | 0 | 0 | 0 | 0 | 0 | 0 | 0 | 100% |
| Boot Fac2 | 0 | 100 | 0 | 0 | 0 | 0 | 0 | 0 | 0 | 0 | 100% |
| Boot Fac3 | 0 | 1 | 97 | 0 | 0 | 0 | 0 | 0 | 0 | 2 | 97% |
| Boot Fac4 | 0 | 2 | 0 | 86 | 0 | 0 | 2 | 0 | 0 | 10 | 86% |
| Boot Fac5 | 1 | 4 | 0 | 0 | 82 | 0 | 0 | 0 | 0 | 13 | 82% |
| Boot Fac6 | 1 | 0 | 0 | 0 | 0 | 99 | 0 | 0 | 0 | 0 | 99% |
| Boot Fac7 | 0 | 0 | 0 | 0 | 0 | 0 | 100 | 0 | 0 | 0 | 100% |
| Boot Fac8 | 0 | 0 | 0 | 0 | 0 | 0 | 0 | 100 | 0 | 0 | 100% |
| Boot Fac9 | 0 | 0 | 0 | 0 | 0 | 0 | 0 | 0 | 100 | 0 | 100% |

**2) Rotational ambiguity**

We first looked at the G-space plot (graphical method) to explore possible rotational ambiguity. As shown in **Figure S3**, there are no obvious oblique edges (to form a V-shaped datapoint distribution) in all the G-space plots. In addition, we did the displacement (DISP) analysis (quantitative method). We found no factor swaps in the DISP analysis with the constraint of d$Q$ smaller than 4, 8, 16, and 32. Therefore, we concluded that there was no significant rotational ambiguity in our PMF solutions. In **Figure S4**, we also present the DISP intervals of all pollutants in all factor profiles.

**Figure S3.** G-space plot for 9 factors in the constrained PMF solution.

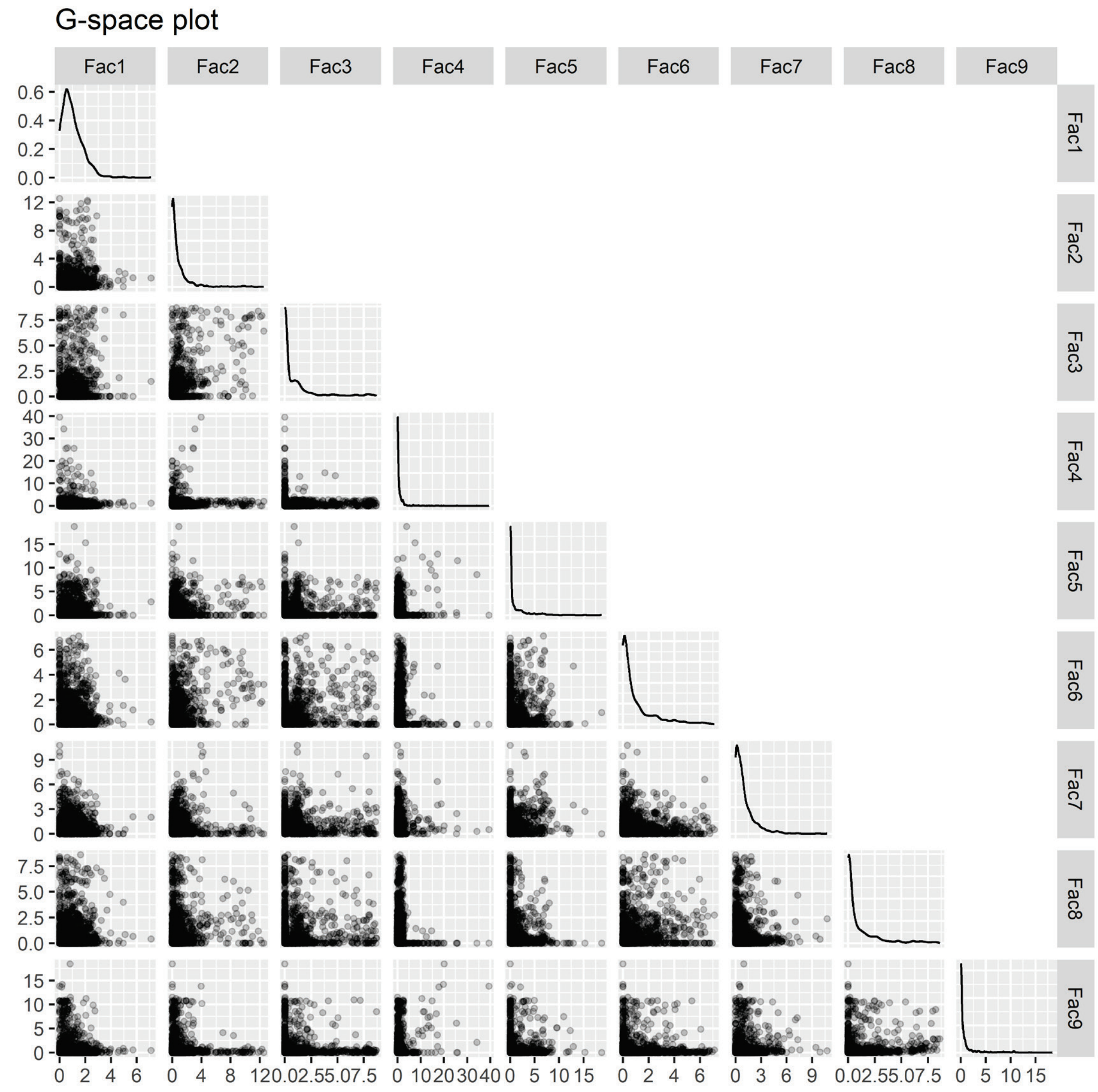

**Figure S4.** Profile of the nine PMF factors with DISP intervals. Dark and light blue refers to strong and weak species, respectively.

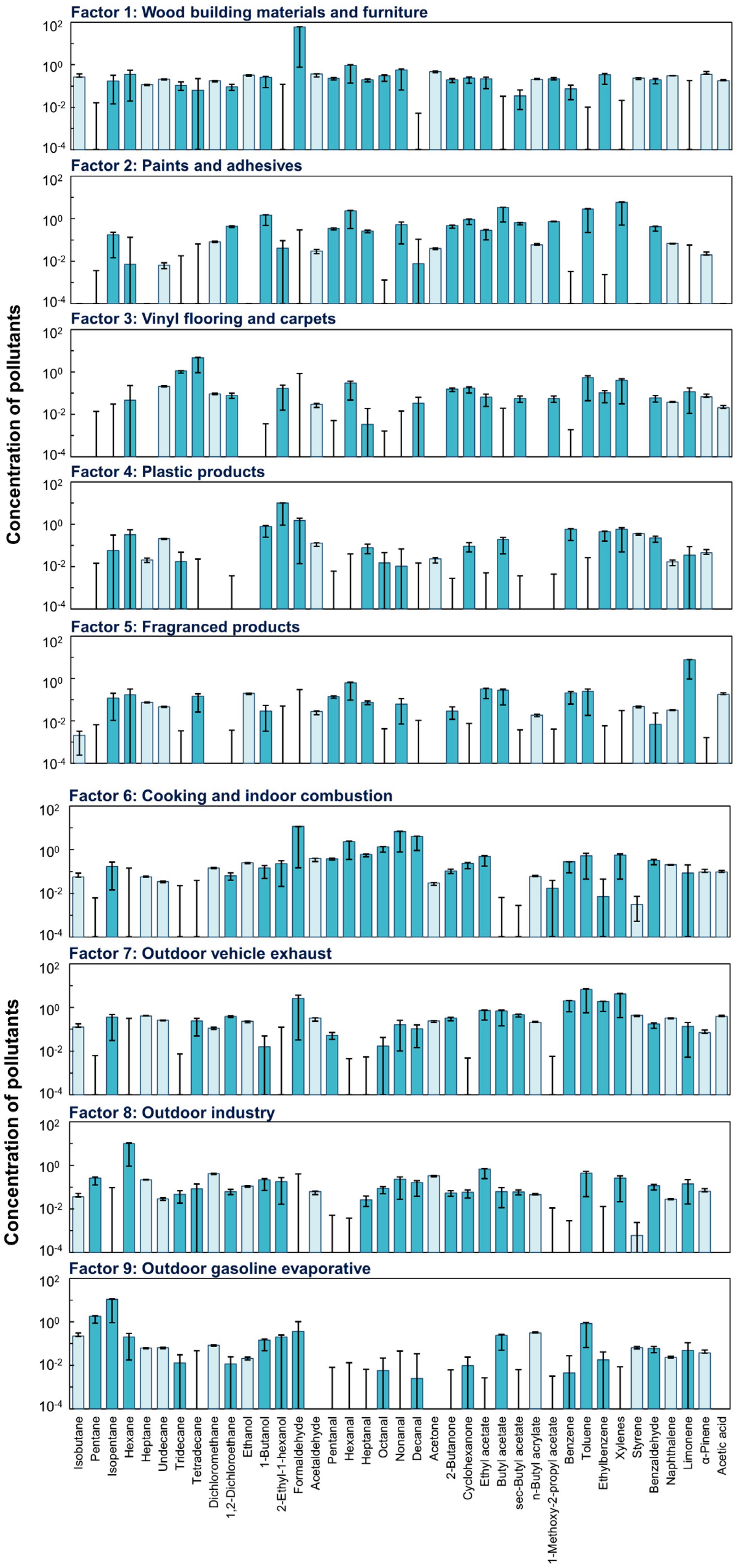

### Section S3 Burden of disease estimation methods

#### S3.1 Exposure-response (E-R) relationships

Two types of E-R relationships were included in this study. The E-R relationships for formaldehyde, benzene, and toluene were obtained from previous meta-analyses based on available epidemiological studies. [13, 14] These relationships were based on relative risks (RR), which is shown as below. [13, 14]

$$RR = \begin{cases} RR_0^{\frac{C}{C_0}}, C \le C_{\max} \\ RR_0^{\frac{C_{\max}}{C_0}}, C > C_{\max} \end{cases} \quad \text{(S2)}$$

where $RR$ is the relative risk at concentration $C$; $RR_0$ is the relative risk per unit increase of concentration; $C$ is the evaluated concentration; $C_0$ is the unit increase of concentration used in the E-R relationship; and $C_{\max}$ is the maximum level of the E-R curve reported in the literature. The parameters of $RR_0$, $C_0$, and $C_{\max}$ can be found in **Table S3**.

Besides, the inhalation unit risks (UR) of cancers were collected for formaldehyde, dichloromethane, 1,2-dichloroethane, acetaldehyde, ethylbenzene, styrene, and naphthalene from U.S. Environmental Protection Agency (EPA), California EPA, and Michigan Department of Environment, Great Lakes, and Energy (EGLE). [15-17] Then the E-R relationships are shown as below.

$$LCR = C \times UR \quad \text{(S3)}$$

where $LCR$ is the lifetime cancer risk; and $UR$ is the inhalation unit risk per 1 μg/m$^3$ increase in concentrations (available in **Table S3**).

**Table S3.** Exposure-response relationships of VOCs.

| Pollutant | Outcome [a] | Metric | Value [b] | Unit | $C_{max}$ | Source |
|---|---|---|---|---|---|---|
| Formaldehyde | Asthma (Adults) | RR | 1.09 (1.03 – 1.15) | 10 μg/m$^3$ | 97.4 μg/m$^3$ | Meta-analysis [14] |
| | Asthma (Children) | RR | 1.27 (1.20 – 1.35) | 10 μg/m$^3$ | 214.0 μg/m$^3$ | Meta-analysis [14] |
| | Nasopharynx cancer, leukemia, other malignant neoplasms | UR | $1.30\times10^{-5}$ | 1 μg/m$^3$ | —— | US EPA [17] |
| Benzene | Asthma (Children) | RR | 1.08 (1.02 – 1.14) | 1 μg/m$^3$ | 35.1 μg/m$^3$ | Meta-analysis [13] |
| | Leukemia | RR | 1.10 (1.05 – 1.15) | 1 μg/m$^3$ | 12.0 μg/m$^3$ | Meta-analysis [13] |
| Toluene | Asthma | RR | 1.02 (1.00 – 1.04) | 1 μg/m$^3$ | 44.5 μg/m$^3$ | Meta-analysis [13] |
| Dichloromethane | Lung cancer, liver cancer | UR | $1.00\times10^{-8}$ | 1 μg/m$^3$ | —— | US EPA [17] |
| 1,2-Dichloroethane | Soft tissue sarcomas | UR | $2.60\times10^{-5}$ | 1 μg/m$^3$ | —— | US EPA [17] |
| Acetaldehyde | Non-melanoma skin cancer | UR | $2.20\times10^{-6}$ | 1 μg/m$^3$ | —— | US EPA [17] |
| Ethylbenzene | Kidney cancer | UR | $2.50\times10^{-6}$ | 1 μg/m$^3$ | —— | Cal EPA [15] |
| Styrene | Leukemia, Non-Hodgkin lymphoma | UR | $5.70\times10^{-7}$ | 1 μg/m$^3$ | —— | EGLE [16] |
| Naphthalene | Other malignant neoplasms | UR | $3.40\times10^{-5}$ | 1 μg/m$^3$ | —— | Cal EPA [15] |

[a] The name of health outcome is consistent with those used in the GBD study. [18]

[b] If the metric is RR, the value is $RR_0$ with its 95% uncertainty interval. If the metric is UR, the value is the 95% upper limit of $UR$. The EPA databases do not provide other summary statistics for URs.

#### S3.2 Total DALYs, baseline incidence and other data

The province-level total DALYs in China were available in 2017, as reported by Zhou et al. [19] However, due to model changes from GBD 2017 to GBD 2019 and 2021, there is a slight difference in the estimated total DALYs. From the GBD database, we can only acquire the national-level total DALYs for China in 2017. Therefore, the final estimate of DALY for outcome $d$ in province $c$ in 2017 (denoted as $DALY_{total,d,c,adjusted}$) should be adjusted as

$$DALY_{total,d,c,adjusted} = DALY_{total,d,c} \times \frac{DALY_{total,d,China}}{\sum_{c} DALY_{total,d,c}} \quad \text{(S4)}$$

where $DALY_{total,d,c}$ is the total DALY for outcome $d$ in province $c$ in 2017 from Zhou et al.; [19] $DALY_{total,d,China}$ is the national-level total DALY for outcome $d$ in China in 2017 provided by the GBD database. [18] The final estimated results of total DALYs for the included health outcomes (i.e., asthma, nasopharynx cancer, lung cancer, liver cancer, kidney cancer, soft tissue sarcomas, non-melanoma skin cancer, non-Hodgkin lymphoma, leukemia, and other malignant neoplasms) are listed in **Table S8** in **Supporting Information (SI) 2**. These DALYs provide critical data for Equation (9) in the main text.

For estimating the attributable burden of disease based on URs, it is necessary to acquire the baseline incidence data for cancers. The baseline cancer incidence (i.e., the number of cancer cases) in the included 9 provinces in China in 2017 was obtained from World Health Organization (WHO) Cancer Incidence in Five Continents Database Volume XII (CI5-XII) as well as China Cancer Registry Annual Report. [20, 21] Detailed data can be found in **Table S8**.

In addition to total DALYs and baseline incidence data, the estimation of source-specific burden of disease needs population data, life expectancy data, and GDP data in each province in China (**Table S8**), which were obtained from National Bureau of Statistics in China. [22]

#### S3.3 Uncertainty analysis

The uncertainty analysis method of the source contributions and attributable DALYs has been briefly introduced in the main text. This section provides more details for the two-stage Monte Carlo simulation. [23-25]

**Table S4**. The process of a two-stage Monte Carlo simulation.

| | $C_1$ | $C_2$ | …… | $C_{3000}$ | $\overline{RR}$ | $PAF$ | $P_k$ | $PAF_k$ |
|---|---|---|---|---|---|---|---|---|
| $\boldsymbol{RR_{0,1}}$ | $RR_{1,1}$ | $RR_{1,2}$ | …… | $RR_{1,3000}$ | $\overline{RR_1}$ | $PAF_1$ | $P_{k,1}$ | $PAF_{k,1}$ |
| $\boldsymbol{RR_{0,2}}$ | $RR_{2,1}$ | $RR_{2,2}$ | …… | $RR_{2,3000}$ | $\overline{RR_2}$ | $PAF_2$ | $P_{k,2}$ | $PAF_{k,2}$ |
| **……** | …… | …… | …… | …… | …… | …… | …… | …… |
| $\boldsymbol{RR_{0,3000}}$ | $RR_{3000,1}$ | $RR_{3000,2}$ | …… | $RR_{3000,3000}$ | $\overline{RR_{3000}}$ | $PAF_{3000}$ | $P_{k,3000}$ | $PAF_{k,3000}$ |

The first stage reflects the pollutant concentration distribution due to intrapopulation variation. In this stage, for formaldehyde, benzene, and toluene modeled in Liu et al., [24] 3000 concentrations were randomly sampled from the log-normal distributed VOC concentrations from that paper, i.e., $C_1$, ……, $C_{3000}$. For dichloromethane, 1,2-dichloroethane, acetaldehyde, ethylbenzene, styrene, and naphthalene, 3000 concentrations were obtained by bootstrapping the original measured concentrations.

The second stage shows the uncertainties in the exposure-response relationships as well as the source contribution percentage. In this stage, 3000 relative risks per unit increase of VOC concentration ($RR_0$ from the meta-analysis) were randomly sampled from the log-normal distribution of $RR_0$ (log $RR_0$ is approximately normally distributed when sample size is large), i.e., $RR_{0,1}$, ……, $RR_{0,3000}$ in **Table S4**. According to the exposure-response relationship in Equation (S2), a total of 9,000,000 relative risks can be obtained ($RR_{i,j}$ for the $i^{th}$ $RR_0$ and the $j^{th}$ concentration), as is shown in **Table S4**. If URs were used for the exposure-response relationship, due to lack of other distribution information, 3000 identical URs were used to replace $RR_{0,1}$, ……, $RR_{0,3000}$ in **Table S4**.

Then, we averaged each row of $RR_{i,j}$ to obtain 3000 average relative risks, so as to average the variability and retain the uncertainty in the exposure-response relationships. Then the PAF can be estimated for each row, which gives a total of 3000 PAFs. The 100 block bootstrap runs in PMF modeling (**Section S2.2**) can provide 100 random results for the average source contribution percentage (i.e., $P_k$ in **Table S4** and $P_{jk,c}$ in Equations (5) and (8) in the main text). However, there are a few bootstrap runs which generate factor profiles unmapped to any of the base profiles. In order to guarantee the interpretability of factor profiles, we removed all bootstrap runs with unmapped factors and finally remained 67 bootstrap runs. Then we bootstrapped these 67 bootstrap results to obtain 3000 values, i.e., $P_{k,1}$, ……, $P_{k,3000}$. Using these 3000 PAFs and 3000 $P_k$s, the 3000 attributable DALYs and 3000 financial costs can be estimated, which formed the distribution of these values. Finally, the 2.5$^{th}$ to 97.5$^{th}$ percentiles of the 3000 simulation results are reported as the 95% uncertainty intervals of their values.

### Section S4 Source-specific exposure assessment

#### S4.1 Source apportionment

The main results and factor interpretation of source apportionment were presented in Section 3.1 in the main text. This section provides the detailed factor profiles in **Table S5**, as well as the association between factor contribution and external variables in **Figures S5-S7**.

**Table S5.** Factor profiles (pollutant concentrations in each factor, μg/m$^3$) of PMF results.

| Pollutant | Fac1 | Fac2 | Fac3 | Fac4 | Fac5 | Fac6 | Fac7 | Fac8 | Fac9 |
|---|---|---|---|---|---|---|---|---|---|
| Isobutane | 0.27 | 0.00 | 0.00 | 0.00 | 0.00 | 0.06 | 0.13 | 0.04 | 0.22 |
| Pentane | 0.00 | 0.00 | 0.00 | 0.00 | 0.00 | 0.00 | 0.00 | 0.26 | 1.80 |
| Isopentane | 0.17 | 0.17 | 0.00 | 0.06 | 0.12 | 0.17 | 0.36 | 0.00 | 10.73 |
| Hexane | 0.36 | 0.01 | 0.05 | 0.32 | 0.18 | 0.00 | 0.00 | 9.98 | 0.20 |
| Heptane | 0.11 | 0.00 | 0.00 | 0.02 | 0.08 | 0.06 | 0.41 | 0.22 | 0.06 |
| Undecane | 0.21 | 0.01 | 0.21 | 0.21 | 0.05 | 0.03 | 0.27 | 0.03 | 0.07 |
| Tridecane | 0.11 | 0.00 | 1.09 | 0.02 | 0.00 | 0.00 | 0.00 | 0.05 | 0.01 |
| Tetradecane | 0.06 | 0.00 | 4.63 | 0.00 | 0.14 | 0.00 | 0.25 | 0.08 | 0.00 |
| Dichloromethane | 0.18 | 0.08 | 0.09 | 0.00 | 0.00 | 0.15 | 0.11 | 0.42 | 0.08 |
| 1,2-Dichloroethane | 0.09 | 0.43 | 0.08 | 0.00 | 0.00 | 0.06 | 0.40 | 0.06 | 0.01 |
| Ethanol | 0.33 | 0.00 | 0.00 | 0.00 | 0.20 | 0.26 | 0.23 | 0.11 | 0.02 |
| 1-Butanol | 0.25 | 1.47 | 0.00 | 0.76 | 0.03 | 0.15 | 0.02 | 0.22 | 0.15 |
| 2-Ethyl-1-hexanol | 0.00 | 0.04 | 0.17 | 9.84 | 0.00 | 0.23 | 0.00 | 0.18 | 0.20 |
| Formaldehyde | 60.41 | 0.00 | 0.00 | 1.49 | 0.00 | 11.71 | 2.60 | 0.00 | 0.36 |
| Acetaldehyde | 0.37 | 0.03 | 0.03 | 0.13 | 0.03 | 0.40 | 0.33 | 0.06 | 0.00 |
| Pentanal | 0.23 | 0.35 | 0.00 | 0.00 | 0.14 | 0.39 | 0.05 | 0.00 | 0.00 |

| **Pollutant** | **Fac1** | **Fac2** | **Fac3** | **Fac4** | **Fac5** | **Fac6** | **Fac7** | **Fac8** | **Fac9** |
|---|---|---|---|---|---|---|---|---|---|
| Hexanal | 0.96 | 2.39 | 0.30 | 0.00 | 0.62 | 2.36 | 0.00 | 0.00 | 0.00 |
| Heptanal | 0.20 | 0.27 | 0.00 | 0.08 | 0.07 | 0.60 | 0.00 | 0.03 | 0.00 |
| Octanal | 0.30 | 0.00 | 0.00 | 0.01 | 0.00 | 1.36 | 0.02 | 0.09 | 0.01 |
| Nonanal | 0.57 | 0.52 | 0.00 | 0.01 | 0.06 | 6.91 | 0.16 | 0.23 | 0.00 |
| Decanal | 0.00 | 0.01 | 0.03 | 0.00 | 0.00 | 4.00 | 0.10 | 0.16 | 0.00 |
| Acetone | 0.48 | 0.04 | 0.00 | 0.02 | 0.00 | 0.03 | 0.24 | 0.33 | 0.00 |
| 2-Butanone | 0.20 | 0.48 | 0.16 | 0.00 | 0.03 | 0.10 | 0.33 | 0.05 | 0.00 |
| Cyclohexanone | 0.24 | 0.92 | 0.17 | 0.09 | 0.00 | 0.23 | 0.00 | 0.06 | 0.01 |
| Ethyl acetate | 0.22 | 0.28 | 0.07 | 0.00 | 0.32 | 0.49 | 0.73 | 0.67 | 0.00 |
| Butyl acetate | 0.00 | 3.40 | 0.00 | 0.19 | 0.27 | 0.00 | 0.71 | 0.06 | 0.24 |
| sec-Butyl acetate | 0.03 | 0.65 | 0.06 | 0.00 | 0.00 | 0.00 | 0.47 | 0.06 | 0.00 |
| n-Butyl acrylate | 0.20 | 0.06 | 0.00 | 0.00 | 0.02 | 0.06 | 0.21 | 0.05 | 0.31 |
| 1-Methoxy-2-propyl acetate | 0.22 | 0.74 | 0.06 | 0.00 | 0.00 | 0.02 | 0.00 | 0.00 | 0.00 |
| Benzene | 0.07 | 0.00 | 0.00 | 0.58 | 0.21 | 0.28 | 2.05 | 0.00 | 0.00 |
| Toluene | 0.00 | 2.77 | 0.54 | 0.00 | 0.25 | 0.53 | 6.84 | 0.44 | 0.84 |
| Ethylbenzene | 0.34 | 0.00 | 0.10 | 0.44 | 0.00 | 0.01 | 1.85 | 0.00 | 0.02 |
| Xylenes | 0.00 | 5.84 | 0.39 | 0.58 | 0.00 | 0.56 | 4.25 | 0.26 | 0.00 |
| Styrene | 0.23 | 0.00 | 0.00 | 0.36 | 0.05 | 0.00 | 0.44 | 0.00 | 0.07 |
| Benzaldehyde | 0.20 | 0.42 | 0.06 | 0.23 | 0.01 | 0.33 | 0.18 | 0.12 | 0.06 |
| Naphthalene | 0.31 | 0.07 | 0.04 | 0.02 | 0.03 | 0.20 | 0.32 | 0.03 | 0.02 |
| Limonene | 0.00 | 0.00 | 0.12 | 0.03 | 7.69 | 0.08 | 0.14 | 0.14 | 0.05 |
| α-Pinene | 0.36 | 0.02 | 0.07 | 0.05 | 0.00 | 0.09 | 0.07 | 0.06 | 0.04 |
| Acetic acid | 0.18 | 0.00 | 0.02 | 0.00 | 0.18 | 0.09 | 0.39 | 0.00 | 0.00 |
| TVOC | 81.66 | 73.95 | 47.62 | 0.00 | 4.88 | 46.32 | 48.56 | 42.96 | 32.44 |

**Note:** Fac1 – Wood building materials and furniture; Fac2 – Paints and adhesives; Fac3 – Vinyl flooring and carpets; Fac4 – Plastic products; Fac5 – Fragranced products; Fac6 – Cooking and indoor combustion; Fac7 – Outdoor vehicle exhaust; Fac8 – Outdoor industry; Fac9 – Outdoor gasoline evaporative.

**Figure S5.** Difference in factor contributions between summer and winter.

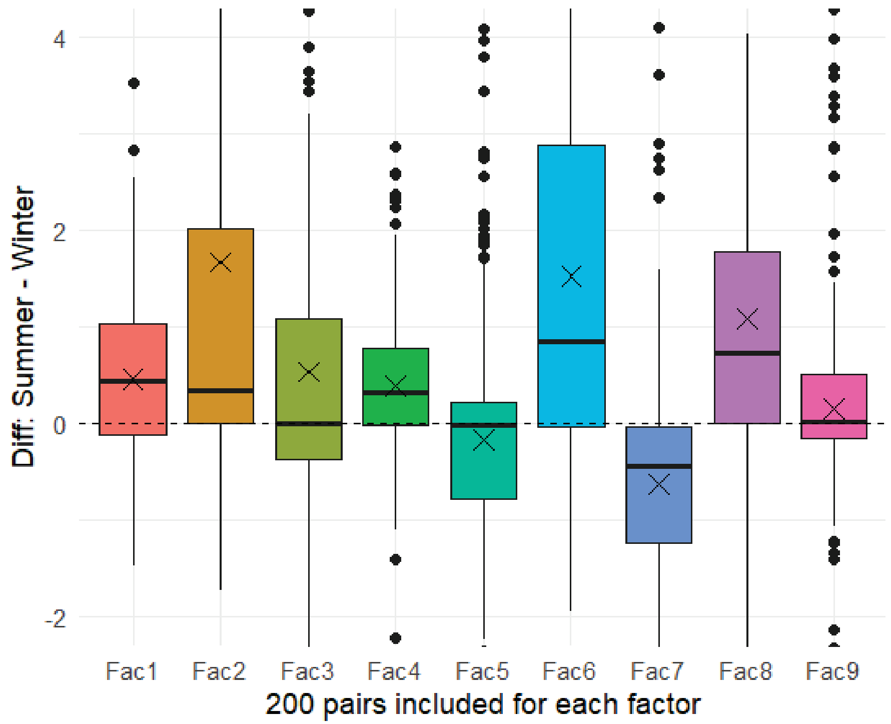

**Figure S6.** Difference in factor contributions between kitchen and non-kitchen.

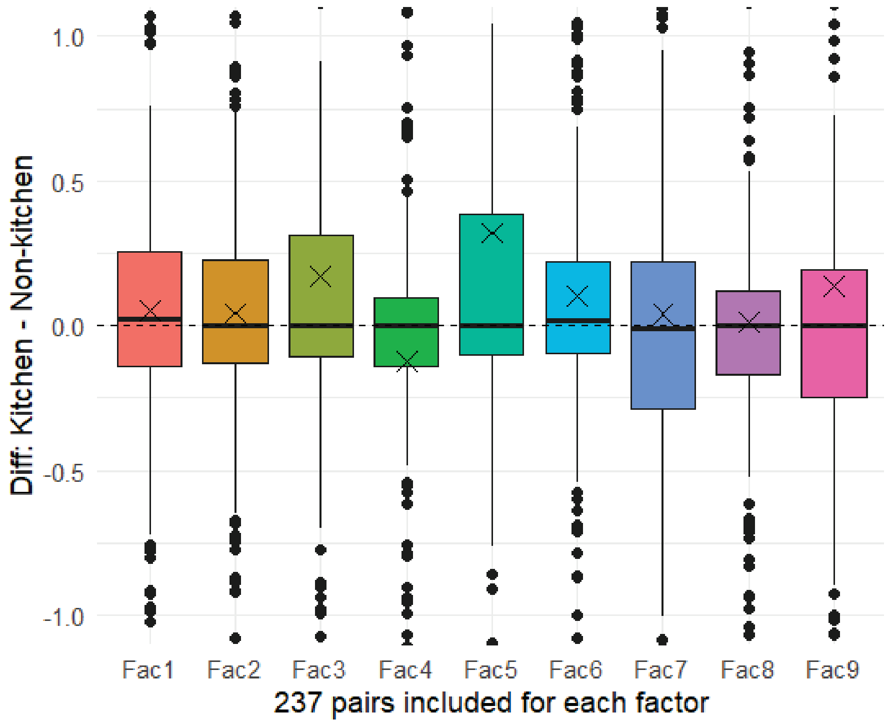


**Figure S7.** Factor contributions under different renovation times. The number at the bottom refers to the sample size in each subgroup stratified by the renovation time.

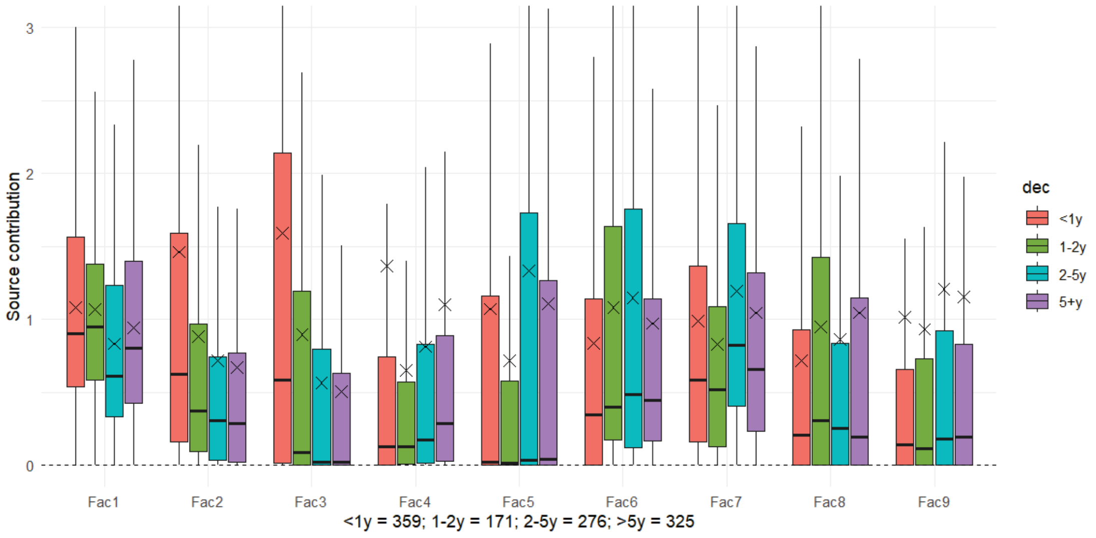

#### S4.2 Source-specific contribution to VOC concentrations

First of all, summary statistics of residential concentrations of 39 included VOCs and TVOCs are shown in **Table S9** in SI2 and briefly discussed in Section 3.2 in the main text.

Then, the source-specific contribution to TVOC concentrations has been discussed in Section 3.2 in the main text. The source-specific contribution to concentrations of 39 included VOCs is shown in **Table S10** in SI2. The ranking of different sources for each included province according to their contribution to TVOC concentrations is shown in **Table S11** in SI2.

Besides TVOCs, the source contribution to concentrations of different specific VOCs could be quite different. The following **Figures S8-S10** provide some representative examples, including formaldehyde, benzene, and 1,2-dichloroethane.

**Figure S8.** Proportion of source contribution to formaldehyde concentration.

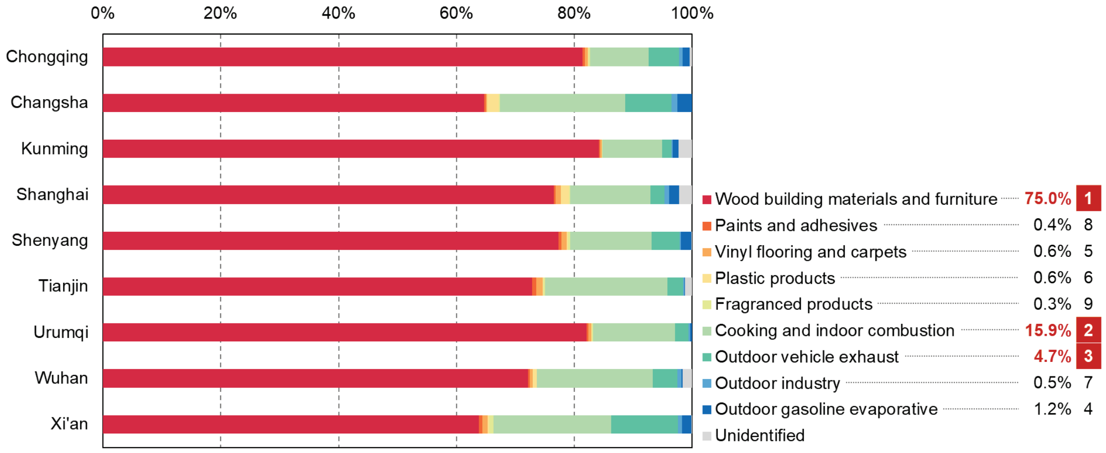

**Figure S9.** Proportion of source contribution to benzene concentration.

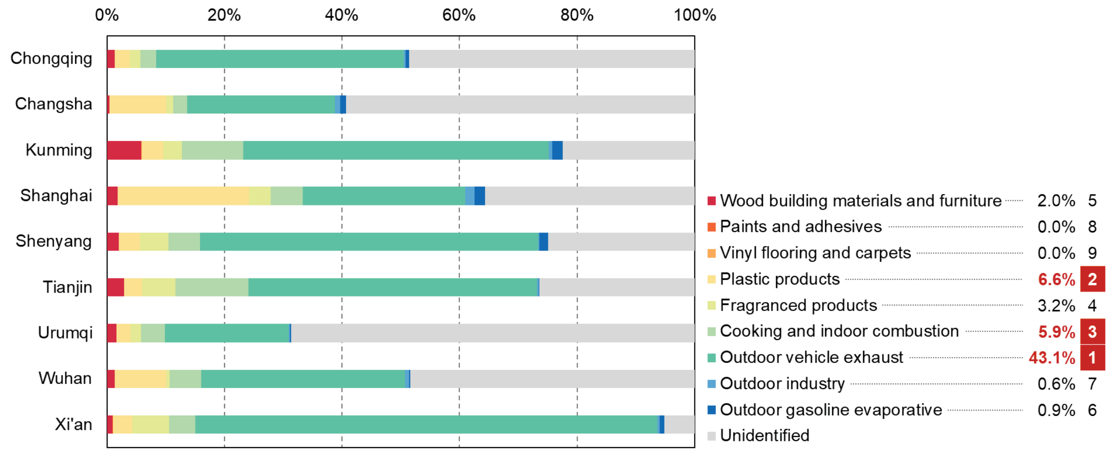


**Figure S10.** Proportion of source contribution to hexanal concentration.

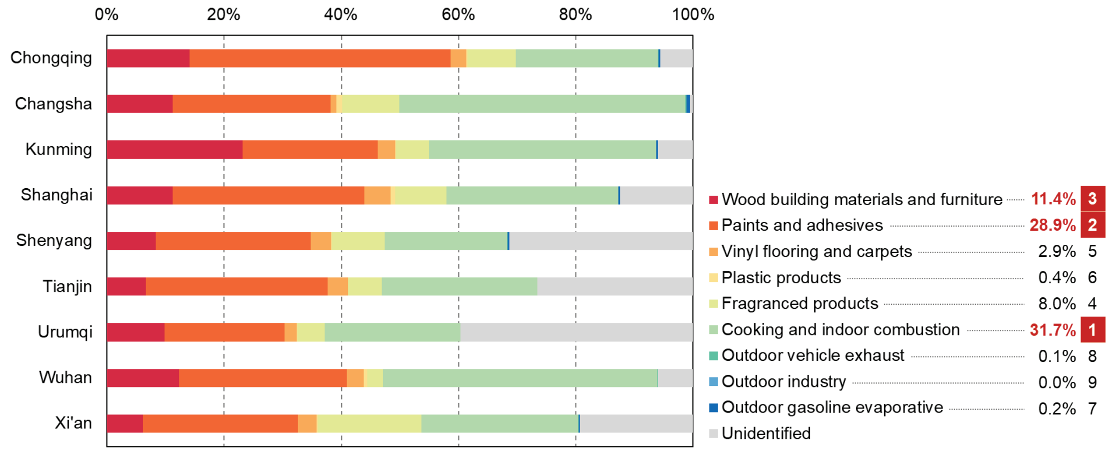

#### S4.3 Multi-room VOC concentration comparison

One of the strengths in this study was measuring the VOC concentrations in multiple rooms (i.e., kitchens, bedrooms, and living rooms) instead of a single room used in previous studies. Therefore, it is necessary to see if significant difference existed in the VOC concentrations among different rooms. If so, then the single-room measurement in previous studies may be biased for indoor exposure assessment.

We built a simple linear mixed model for this analysis. The model is shown below.

$$Conc \sim Room + \left(1 \middle| id\right) \tag{S5}$$

where *Conc* is the concentration of the focused VOC; *Room* is a categorical variable representing kitchens, living rooms, or bedrooms (reference level); each *id* refers to each residence in each season. As people spend the most time in bedrooms at home, we set bedrooms as the reference level. Since there were too many VOCs in this study, here we selected some important VOCs for detailed analysis:

a) Formaldehyde, benzene, toluene: Three VOCs with E-R relationships from the meta-analysis;
b) Acetaldehyde, hexanal, butyl acetate, tetradecane, 2-ethyl-1-hexanol, limonene, hexane, isopentane: tracers for different sources
c) TVOCs

**Table S6.** Regression coefficients of *Room* in different cities for selected VOCs.

| City | Formaldehyde | | Benzene | | Toluene | |
|---|---|---|---|---|---|---|
| | Kitchen | Living | Kitchen | Living | Kitchen | Living |
| Tianjin | 7.391 | -0.516 | -0.119 | -0.207 | -0.606 | -0.494 |
| Urumqi | 1.015 | -7.851** | -5.845 | -18.375*** | -2.534 | -0.055 |
| Kunming | -4.009 | -9.453** | -1.719* | -1.299 | -1.694 | 6.259 |
| Shenyang | -9.355* | -4.664 | -1.839 | -2.657** | -1.270 | 1.022 |
| Chongqing | -4.181 | -13.482*** | -5.701 | -6.926* | -6.512 | -10.514 |
| Xi’an | 10.503* | -10.217* | 0.840 | -0.535 | 1.722 | 0.994 |
| Shanghai | 3.870 | -7.518 | -5.601* | -5.396* | -10.807** | -10.903** |
| Changsha | -4.100 | -4.388 | -8.319 | -14.285* | -0.468 | -3.284 |
| Wuhan | 8.063 | -3.034 | -4.105 | -1.285 | 0.573 | 2.242 |

| City | Acetaldehyde | | Hexanal | | Butyl acetate | |
|---|---|---|---|---|---|---|
| | Kitchen | Living | Kitchen | Living | Kitchen | Living |
| Tianjin | 0.176 | 0.25 | 2.886 | -1.898 | -3.563 | -3.020 |
| Urumqi | 0.043 | -0.449 | 3.007 | -2.384 | -1.385 | -0.680 |
| Kunming | -0.159 | 0.705 | 1.327 | 0.249 | -0.809 | 0.526 |
| Shenyang | 0.362 | 0.631 | -0.976 | 0.963 | 3.183 | 2.341 |
| Chongqing | -0.756 | -0.908** | -0.698 | -1.857** | -3.253 | -1.510 |
| Xi’an | 0.828 | 1.076** | 8.427** | -1.184 | -0.806 | -1.648 |
| Shanghai | | | | | -4.411 | -4.006 |
| Changsha | | | | | 4.223 | 1.729 |
| Wuhan | | | | -0.716 | -0.838 | 0.952 |

| City | Tetradecane | | 2-ethyl-1-hexanol | | Limonene | |
|---|---|---|---|---|---|---|
| | Kitchen | Living | Kitchen | Living | Kitchen | Living |
| Tianjin | 3.287*** | -0.558 | 0.223 | -0.459 | 3.568 | 2.369 |
| Urumqi | -1.443 | -1.987 | -2.850 | -3.472** | 4.060 | 3.216 |
| Kunming | 0.271 | -0.185 | -0.418 | 1.217 | 18.218 | 0.143 |
| Shenyang | 0.396 | 1.128 | -0.830 | 2.689 | -10.118* | -4.053 |
| Chongqing | -0.561 | -2.308** | -1.650 | -1.965 | -1.328 | 0.731 |
| Xi’an | 1.530 | -1.743 | -1.323 | 0.638 | 45.432*** | 10.727 |
| Shanghai | | | | | | |
| Changsha | | | | | | |
| Wuhan | | -0.810 | | 2.167 | | 0.754 |

| City | Hexane | | Isopentane | | TVOCs | |
|---|---|---|---|---|---|---|
| | Kitchen | Living | Kitchen | Living | Kitchen | Living |
| Tianjin | -1.199 | 3.660* | 0.787 | 4.360 | -13.7 | -69.4* |
| Urumqi | 4.731 | 5.841* | -0.226 | 0.143 | -93.1 | -134.4* |
| Kunming | -0.918 | 2.173 | -6.912 | -5.889 | 10.0 | 26.4 |
| Shenyang | -3.787 | -2.008 | 76.002 | -29.42 | -302.6 | -319.0 |
| Chongqing | -32.703 | -29.306** | 24.453** | 7.179 | -120.6 | -170.5* |
| Xi’an | 8.451 | 19.994 | -4.451 | -3.091 | 20.6 | 91.6 |
| Shanghai | | | | | -74.0 | -87.9* |
| Changsha | | | | | -301.4 | -330.8* |
| Wuhan | | 5.803 | | 1.095 | -198.5** | -121.6* |

Note: * $p < 0.05$, ** $p < 0.01$, *** $p < 0.001$; Bedrooms as reference level.

As can be seen from **Table S6**, there were significant differences in concentrations among different rooms, especially for living rooms and for formaldehyde, benzene, and TVOCs. For other selected VOCs, there was no significant concentration difference in most cities. The VOC concentrations in living rooms were usually significantly lower than those in bedrooms, possibly because the bedrooms usually had a higher loading ratio of the emission sources (i.e., higher emission area per volume of room) and a lower ventilation rate. The finding suggests that collecting VOC data in a single room was not enough to characterize the indoor concentrations. Future studies should measure VOC concentrations in multiple rooms.

#### S4.4 PMF model performance for different cities

Although the PMF results have been validated to be reliable and robust, the assumption of spatiotemporal constant profile in this study may result in model underfit in some cities. We calculated the explained proportion of TVOC concentrations by the PMF model, which was 79.7%, 74.7%, 80.2%, 77.3%, 61.4%, 90.4%, 72.0%, 90.4%, and 87.3% for Chongqing, Changsha, Kunming, Shanghai, Shenyang, Tianjin, Urumqi, Wuhan, and Xi'an in China. The model performance for **Shenyang** (61.4%) was relatively poor compared to other cities (mostly higher than 75%). If we checked the VOC-specific explained proportion in Shenyang, the poor fit (<70%) was observed for pentane (58%), hexanal (66%), and toluene (65%). Compared with other cities, Shenyang is located in northeast China with severe cold climate, which can influence the window-opening behavior of residents and contributions from central heating sources. Shenyang is also one of the most famous heavy industry base in China. The traditional heavy industry can also provide special industrial emission sources, which were not identified by the current PMF model and could explain more about pentane and toluene. Besides, the model fit for **Urumqi** (72.0%) was also not so ideal, with poor performance in isopentane (66%), 1-butanol (55%), hexanal (59%), butyl acetate (65%), PGMEA (53%), benzene (30%), and toluene (59%). Urumqi is one of the most important cities in northwest China with severe cold but very dry weather. It is also populated by various ethnic groups like Uyghur, who usually have different lifestyles and habits, resulting in different characteristics of indoor sources. The species with low explained percentage seems to suggest that there were other unidentified solvent-related indoor sources. More data need to be collected in the future such as questionnaire data to explore the features of residential VOC sources in different cities.

#### Section S5 Source-specific burden of disease

The detailed results of source-specific burden of disease attributable to residential VOCs are presented in **Tables S12-S14** in SI2, including DALY number, rate, and proportion. The corresponding financial costs can be found in **Tables S15-S17** in SI2, including total costs, cost per capita, and proportion of GDP. The source-specific health burden for specific health outcomes is available in **Tables S18-S19** in SI2. Finally, the ranking of sources according to health burden for each province is in **Table S20** in SI2.

**Figure S11.** Provincial-level DALY rates attributable to VOCs in China's residences, by source, 2017.

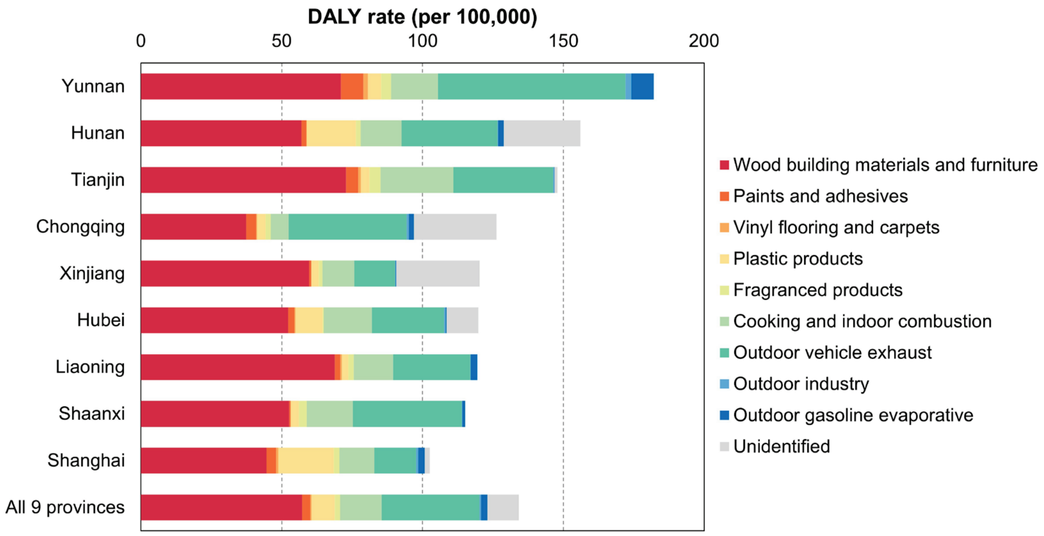

**Figure S12.** Spatial distribution of attributable DALY proportion of VOCs from top three sources in 2017.

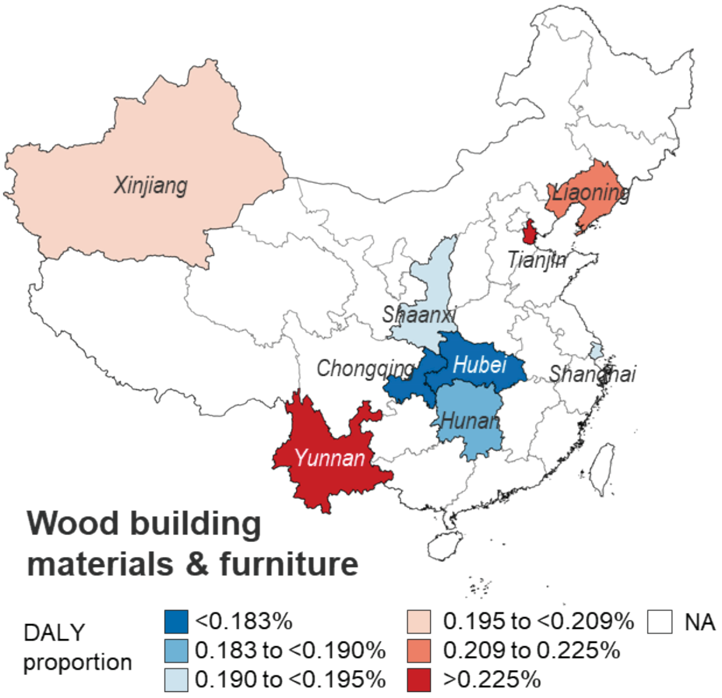


(*a*) Wood building materials and furniture

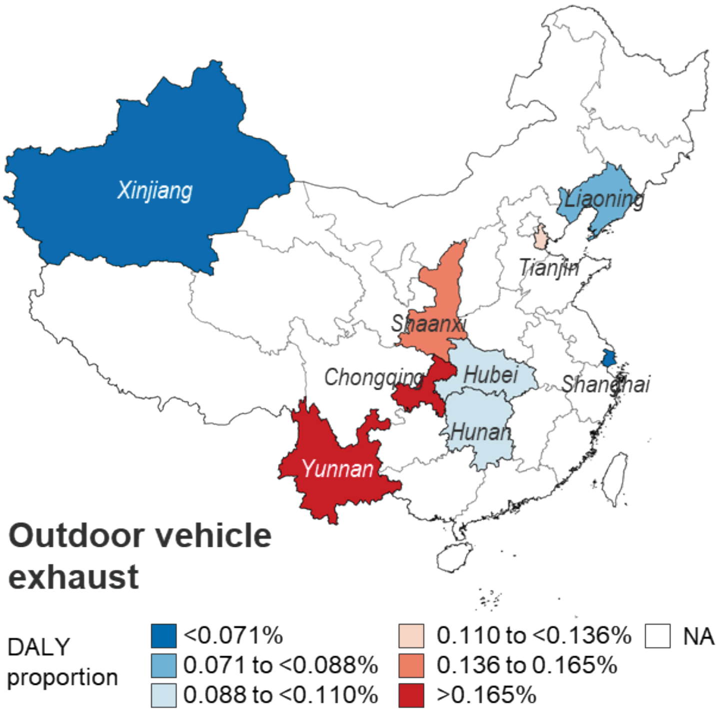


(*b*) Outdoor vehicle exhaust

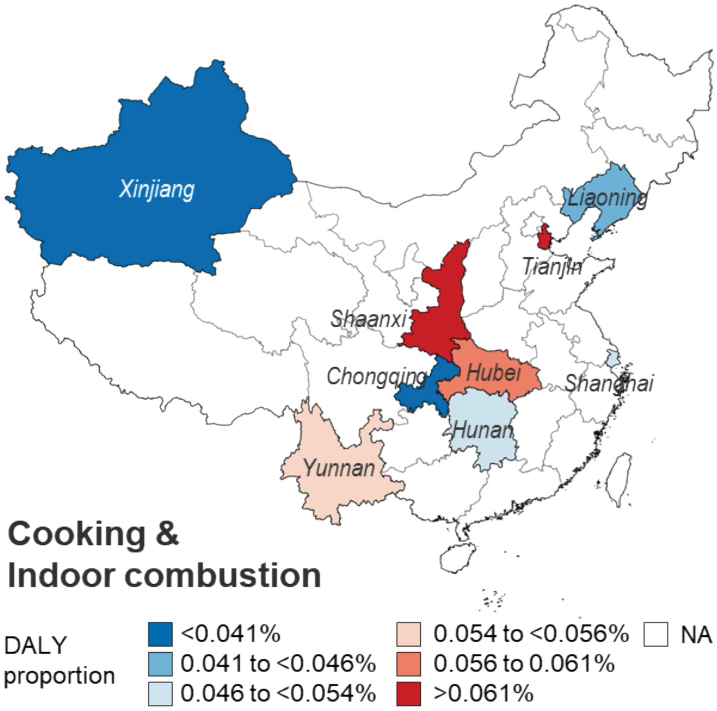


(*c*) Cooking and indoor combustion

**Figure S13.** Point estimates and 95% UIs of source-specific VOC-attributable DALYs by outcome in China in 2017.

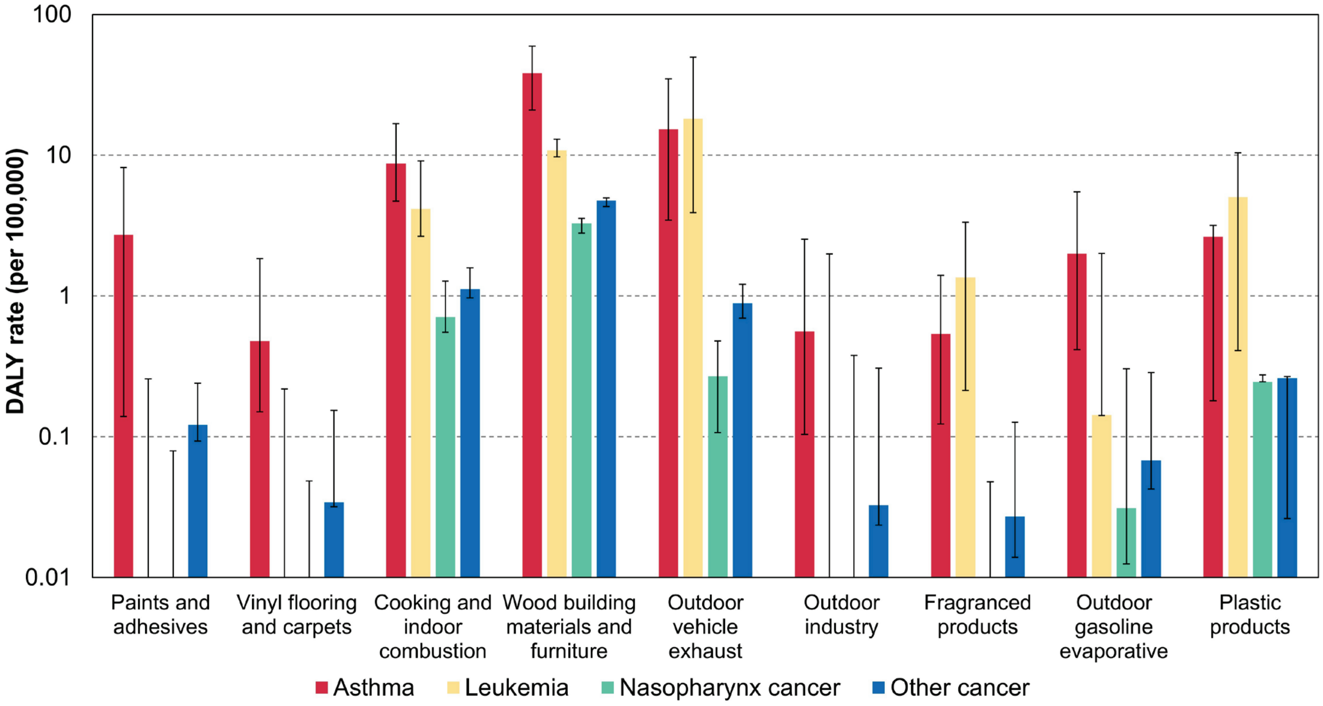


**Note:** To show the relatively low results for some outcomes, the y-axis is transformed into the logarithm scale. Since the cancer unit risks from US EPA only had the point estimate without any uncertainty intervals, so the 95% UIs of nasopharynx cancer and other cancer only considered the uncertainties in PMF results. For leukemia, the uncertainty originated from both PMF and E-R relationships for benzene, but only PMF for formaldehyde and styrene. The 95% UIs of asthma considered both two sources of uncertainties.

#### Section S6 Comparison with previous studies

**Table S7.** Comparison with previous studies about source contributions to TVOC concentrations.

| Source | This study (9 cities, China) [a] | Xi'an, China [b] [26] | Beijing, China [c] [27] | Edmonton, Canada [d] [28] |
|---|---|---|---|---|
| Building materials and furniture | 30.3% | 44.5% | 31.8% | 8.0% |
| Paints and adhesives | 16.6% | 11.9% | 16.4% | 3.0% |
| Household consumer products | 4.0% | 17.3% | —— | 58.3% |
| Cooking and indoor combustion | 13.6% | 24.3% | 24.4% | 10.5% |
| Outdoor vehicle | 25.1% | 2.1% | 6.0% | 9.9% |
| Outdoor industry | 10.5% | —— | 10.2% | 10.3% |

[a] The percentage is scaled to make the sum equal to 100%, which is consistent with other columns. Building materials and furniture combines factors 1 and 3; Household consumer products combines factors 4 and 5; Outdoor vehicle combines factors 7 and 9.

[b] Cooking and indoor combustion combines cooking and smoking.

[c] We used the source contributions to each VOC and the weighted average method to estimate the source contributions to TVOC concentrations for this study. Building materials and furniture combines wooden flooring and wooden furniture; Paints and adhesives uses wall coverings; Cooking and indoor combustion uses ozone-initiated reactions; Outdoor industry uses halogenated hydrocarbons of miscellaneous outdoor origin.

[d] Building materials and furniture combines building materials and floor and wall coverings; Household consumer products combines household products, fragranced consumer products, deodorizers, and tap water and bleach use; Outdoor vehicle combines traffic emissions, fuel evaporation, gasoline production and storage, and industrial and vehicular evaporative; Outdoor industry uses oil and gas industry.

### Reference


1. U.S. Environmental Protection Agency *EPA Positive Matrix Factorization (PMF) 5.0 Fundamentals and User Guide*; 2014.

2. Liu, N.; Oshan, R.; Blanco, M.; Sheppard, L.; Seto, E.; Larson, T.; Austin, E., Mapping Source-Specific Air Pollution Exposures Using Positive Matrix Factorization Applied to Multipollutant Mobile Monitoring in Seattle, WA. *Environ Sci Technol* **2025,** *59*, (7), 3443-3458.

3. Lee, E.; Chan, C. K.; Paatero, P., Application of positive matrix factorization in source apportionment of particulate pollutants in Hong Kong. *Atmos Environ* **1999,** *33*, (19), 3201-3212.

4. Baudic, A.; Gros, V.; Sauvage, S.; Locoge, N.; Sanchez, O.; Sarda-Estève, R.; Kalogridis, C.; Petit, J. E.; Bonnaire, N.; Baisnée, D.; Favez, O.; Albinet, A.; Sciare, J.; Bonsang, B., Seasonal variability and source apportionment of volatile organic compounds (VOCs) in the Paris megacity (France). *Atmos Chem Phys* **2016,** *16*, (18), 11961-11989.

5. Liao, H. T.; Yau, Y. C.; Huang, C. S.; Chen, N.; Chow, J. C.; Watson, J. G.; Tsai, S. W.; Chou, C. C. K.; Wu, C. F., Source apportionment of urban air pollutants using constrained receptor models with a priori profile information. *Environ Pollut* **2017,** *227*, 323-333.

6. Wang, M.; Wang, Q. Y.; Ho, S. S. H.; Li, H.; Zhang, R. J.; Ran, W. K.; Qu, L. L.; Lee, S. C.; Cao, J. J., Chemical characteristics and sources of nitrogen-containing organic compounds at a regional site in the North China Plain during the transition period of autumn and winter. *Sci Total Environ* **2022,** *812*, 151451.

7. Huang, C. S.; Liu, Y. H.; Liao, H. T.; Chen, C. Y.; Wu, C. F., Improvements in source apportionment of multiple time-resolved PM(2.5) inorganic and organic speciation measurements using constrained Positive Matrix Factorization. *Environ Sci Pollut Res Int* **2024,** *31*, (55), 64185-64198.

8. Huang, C. S.; Liao, H. T.; Lu, S. H.; Chan, C. C.; Wu, C. F., Identifying and quantifying PM(2.5) pollution episodes with a fusion method of moving window technique and constrained Positive Matrix Factorization. *Environ Pollut* **2022,** *315*, 120382.

9. Huang, Y.; Ho, S. S. H.; Ho, K. F.; Lee, S. C.; Yu, J. Z.; Louie, P. K. K., Characteristics and health impacts of VOCs and carbonyls associated with residential cooking activities in Hong Kong. *J Hazard Mater* **2011,** *186*, (1), 344-351.

10. Li, L. X.; Cheng, Y.; Dai, Q. L.; Liu, B. S.; Wu, J. H.; Bi, X. H.; Choe, T. H.; Feng, Y. C., Chemical characterization and health risk assessment of VOCs and PM2.5-bound

PAHs emitted from typical Chinese residential cooking. *Atmos Environ* **2022,** *291*.

11. Huang, L.; Cheng, H. N.; Ma, S. T.; He, R. Y.; Gong, J. C.; Li, G. Y.; An, T. C., The exposures and health effects of benzene, toluene and naphthalene for Chinese chefs in multiple cooking styles of kitchens. *Environ Int* **2021,** *156*.

12. Zheng, Z. H.; Zhang, H. M.; Qian, H.; Li, J. G.; Yu, T.; Liu, C., Emission characteristics of formaldehyde from natural gas combustion and effects of hood exhaust in Chinese kitchens. *Sci Total Environ* **2022,** *838*.

13. Liu, N. R.; Bu, Z. M.; Liu, W.; Kan, H. D.; Zhao, Z. H.; Deng, F. R.; Huang, C.; Zhao, B.; Zeng, X. G.; Sun, Y. X.; Qian, H.; Mo, J. H.; Sun, C. J.; Guo, J. G.; Zheng, X. H.; Weschler, L. B.; Zhang, Y. P., Health effects of exposure to indoor volatile organic compounds from 1980 to 2017: A systematic review and meta-analysis. *Indoor Air* **2022,** *32*, (5), e13038.

14. Liu, N. R.; Fang, L.; Liu, W.; Kan, H. D.; Zhao, Z. H.; Deng, F. R.; Huang, C.; Zhao, B.; Zeng, X. A.; Sun, Y. X.; Qian, H.; Mo, J. H.; Sun, C. J.; Guo, J. G.; Zheng, X. H.; Bu, Z. M.; Weschler, L. B.; Zhang, Y. P., Health effects of exposure to indoor formaldehyde in civil buildings: A systematic review and meta-analysis on the literature in the past 40 years. *Build Environ* **2023,** *233*, 110080.

15. California Environmental Protection Agency Toxicity criteria on chemicals evaluated by Office of Environmental Health Hazard Assessment (OEHHA). https://oehha.ca.gov/library/chemicals (Apr 8, 2025),

16. Department of Environment, G. L., and Energy in Michigan,, Air Toxics Program. https://www.michigan.gov/egle/about/organization/air-quality/air-toxics (Apr 8, 2025),

17. U.S. Environmental Protection Agency Integrated Risk Information System (IRIS) Assessments. https://iris.epa.gov/AtoZ/?list_type=alpha (Apr 8, 2025),

18. GBD Risk Factors Collaborators, Global burden and strength of evidence for 88 risk factors in 204 countries and 811 subnational locations, 1990-2021: a systematic analysis for the Global Burden of Disease Study 2021. *Lancet* **2024,** *403*, (10440), 2162-2203.

19. Zhou, M. G.; Wang, H. D.; Zeng, X. Y.; Yin, P.; Zhu, J.; Chen, W. Q.; Li, X. H.; Wang, L. J.; Wang, L. M.; Liu, Y. N.; Liu, J. M.; Zhang, M.; Qi, J. L.; Yu, S. C.; Afshin, A.; Gakidou, E.; Glenn, S.; Krish, V. S.; Miller-Petrie, M. K.; Mountjoy-Venning, W. C.; Mullany, E. C.; Redford, S. B.; Liu, H. Y.; Naghavi, M.; Hay, S. I.; Wang, L. H.; Murray, C. J. L.; Liang, X. F., Mortality, morbidity, and risk factors in China and its provinces, 1990-2017: a systematic analysis for the Global Burden of Disease Study 2017. *Lancet* **2019,** *394*, (10204), 1145-1158.

20. He, J.; Wei, W., *China Cancer Registry Annual Report*. People's Medical Publishing House: Beijing, China, 2019.

21. World Health Organization Cancer Incidence in Five Continents Database Volume XII (CI5-XII). https://ci5.iarc.who.int/previous/download (Apr 8, 2025),

22. National Bureau of Statistics in China National data. https://data.stats.gov.cn/easyquery.htm?cn=C01 (Apr 8, 2025),

23. Hu, Y.; Ji, J. S.; Zhao, B., Deaths Attributable to Indoor PM2.5 in Urban China When Outdoor Air Meets 2021 WHO Air Quality Guidelines. *Environ Sci Technol* **2022,** *56*, (22), 15882-15891.

24. Liu, N.; Liu, W.; Deng, F.; Liu, Y.; Gao, X.; Fang, L.; Chen, Z.; Tang, H.; Hong, S.; Pan, M.; Liu, W.; Huo, X.; Guo, K.; Ruan, F.; Zhang, W.; Zhao, B.; Mo, J.; Huang, C.; Su, C.; Sun, C.; Zou, Z.; Li, H.; Sun, Y.; Qian, H.; Zheng, X.; Zeng, X.; Guo, J.; Bu, Z.; Mandin, C.; Hanninen, O.; Ji, J. S.; Weschler, L. B.; Kan, H.; Zhao, Z.; Zhang, Y., The burden of disease attributable to indoor air pollutants in China from 2000 to 2017. *Lancet Planet Health* **2023,** *7*, (11), e900-e911.

25. Liu, Y. M.; Zhou, B.; Wang, J. H.; Zhao, B., Health benefits and cost of using air purifiers to reduce exposure to ambient fine particulate pollution in China. *J Hazard Mater* **2021,** *414*, 125540.

26. Huang, Y.; Su, T.; Wang, L. Q.; Wang, N.; Xue, Y. G.; Dai, W. T.; Lee, S. C.; Cao, J. J.; Ho, S. S. H., Evaluation and characterization of volatile air toxics indoors in a heavy polluted city of northwestern China in wintertime. *Sci Total Environ* **2019,** *662*, 470-480.

27. Huang, L. H.; Qian, H.; Deng, S. X.; Guo, J. F.; Li, Y. P.; Zhao, W. P.; Yue, Y., Urban residential indoor volatile organic compounds in summer, Beijing: Profile, concentration and source characterization. *Atmos Environ* **2018,** *188*, 1-11.

28. Bari, M. A.; Kindzierski, W. B.; Wheeler, A. J.; Héroux, M. E.; Wallace, L. A., Source apportionment of indoor and outdoor volatile organic compounds at homes in Edmonton, Canada. *Build Environ* **2015,** *90*, 114-124.
