## Supplementary material for "Source-specific exposure and burden of disease attributable to volatile organic compounds (VOCs) in China’s residences": Determination letter of UW HSD

### NOT HUMAN SUBJECTS

August 25, 2025

Dear Ningrui Liu:

On 8/25/2025 the University of Washington Human Subjects Division (HSD) reviewed the following application:

| | |
|---|---|
| Type of Review: | Initial Study |
| Title of Study: | Source-specific exposure and burden of disease attributable to volatile organic compounds (VOCs) in China's residences |
| Investigator: | Ningrui Liu |
| IRB ID: | STUDY00023774 |
| Funding: | None<br>Funding Title(s):<br>Pass-through institution(s): |
| IND, IDE, or HDE: | None |

**HSD determined that the proposed activity does not involve human subjects**, as defined by federal and state regulations. Therefore, review and approval by the University of Washington IRB is not required.

This determination applies only to the activities described in this application. **Depending on the nature of your study, you may need to obtain other approvals or permissions to conduct your research. For example, you might need to apply for access to data or specimens (e.g., to obtain UW student data). Or, you might need to obtain permission from facilities managers to conduct activities in the facilities (e.g., Seattle School District; the Harborview Emergency Department).**

HSD does not make determinations on behalf of other institutions. If other institutions are involved in the research, they may need to make their own determination or they may decide to be guided by our determination.

If you need to make changes in the future that may affect this determination or are not sure, contact us or submit a new request for a determination. You can create a modification by clicking Create Modification within the study.

We wish you great success.

Sincerely,

Theresa Naluai-Cecchini
IRB Administrator, Committee J
206.543.3494
